# Chronic intracranial neural dynamics forecast treatment response and therapeutic engagement during deep brain stimulation for OCD

**DOI:** 10.64898/2026.09.18.26363320

**Authors:** Jeffrey Zhou, Rick R. Hanish, Timon Merk, Royal Bao, Sarah Soubra, Anthony K. Allam, Sandesh Reddy, Victoria R. Gates, Vincent Allam, Mark Libowitz, Danika L. Paulo, Kalman A. Katlowitz, Brian J. Mickey, Brent M. Kious, Jeffrey A. Herron, Tomasz M. Frączek, Ben Shofty, Ankit B. Patel, Sarah R. Heilbronner, Eric A. Storch, Wayne K. Goodman, Sameer A. Sheth, Nicole R. Provenza

## Abstract

Clinical improvement following psychiatric neuromodulation often requires weeks to months, creating prolonged uncertainty during treatment optimization. Neural biomarkers could reduce this uncertainty by forecasting treatment outcome, providing early evidence of therapeutic engagement, and objectively tracking clinical response. We analyzed nearly 120,000 patient-hours of chronic intracranial recordings from 24 patients undergoing ventral capsule/ventral striatum deep brain stimulation for treatment-resistant obsessive-compulsive disorder. Before stimulation initiation, circadian neural features distinguished eventual responders from non-responders. Within the first two weeks of therapy, reductions in neural predictability differentiated eventual responders from non-responders and correlated with the magnitude of subsequent symptom improvement, months before stable clinical response was achieved. During chronic treatment, neural predictability tracked clinical response state. Together, these findings establish a framework for neural biomarkers spanning the course of DBS therapy: forecasting treatment response before stimulation, detecting early therapeutic engagement after activation, and longitudinally monitoring clinical response.

## Introduction

Psychiatric disorders are inherently dynamic conditions in which the severity of clinical symptoms continuously evolves over time. On the other hand, clinical evaluations are typically performed intermittently, often weeks or months apart.^1^ These periodic snapshots of the state of the disorder may therefore not fully capture its dynamics. Recent advances in neural recording technologies now enable chronic and continuous monitoring of brain activity, creating new opportunities to relate longitudinal neural dynamics to the clinical trajectory of psychiatric disorders. Neural biomarkers provide a framework for bridging these complementary data streams by linking neural circuit dynamics to clinically meaningful outcomes such as treatment response, symptom severity, or behavior.^2^ Beyond advancing the mechanistic understanding of brain-behavior relationships, objective neural biomarkers could enable more personalized and physiology-guided neuromodulation by informing patient selection, assessing therapeutic target engagement, tracking treatment response, or forecasting clinical outcomes.^3,4^

Sensing-capable deep brain stimulation (DBS) systems provide one such platform, enabling continuous neural recordings directly from the implanted target region throughout the course of therapy.^5,6^ Unlike traditional experimental recordings that capture neural activity only briefly under controlled laboratory conditions, these devices continuously acquire neural signals during patients’ everyday lives while simultaneously delivering therapeutic stimulation.^7^ Biomarkers derived from DBS recordings have demonstrated clinical relevance across multiple neurological and psychiatric disorders, but most have been developed to estimate or track current clinical state, including episodic tic expressions in Tourette syndrome^8–10^ and immediate motor symptoms in Parkinson’s disease,^11–15^ as well as gradual symptom severity changes in depression,^16,17^ post-traumatic stress disorder,^18^ pain,^19^ and obsessive-compulsive disorder (OCD).^20,21^ However, biomarkers capable of forecasting future therapeutic response before clinical improvement becomes apparent remain limited.

The need for such biomarkers is particularly acute in psychiatric DBS, where clinical improvement often requires weeks to months of stimulation.^22–26^ In OCD, therapeutic response after DBS often requires prolonged optimization of stimulation parameters through an iterative trial-and-error process, creating extended periods of uncertainty for both patients and clinicians.^27^ Continuous neural recordings throughout DBS therapy provide the opportunity to determine whether objective neural biomarkers can forecast eventual treatment response before clinical improvement becomes apparent. If successful, such biomarkers could provide an objective neurophysiological measure to guide DBS programming by providing earlier evidence of therapeutic engagement and accelerating treatment optimization.

Emergence of the pathological thoughts and actions observed in OCD is thought to stem from imbalanced activity within the neural circuits governing fear response, reward sensitivity, motivation, controlled decision-making, and related behaviors, including the prefrontal cortex (PFC), ventral striatum (VS), ventral pallidum, thalamus, and extended amygdala.^28–31^ In addition to these gray matter regions, a major component of the circuit is the anterior limb of the internal capsule, a major white matter conduit carrying fibers that interconnect them.^32^ DBS exerts its therapeutic effect by targeting a critical hub within this circuit situated within the ventral portion of the internal capsule (ventral capsule, VC), near its border with the ventral striatum (VS) and bed nucleus of the stria terminalis (BNST^33^, a core component of the extended amygdala). Although many patients derive substantial benefit from DBS in this region, clinical response is heterogeneous and often slow to emerge.^26,27^ Taken together, there is a need for physiological measures that can indicate therapeutic engagement before symptom improvement becomes clinically apparent. Chronic recordings within the VC/VS region therefore provide an opportunity to identify biomarkers of treatment response while sampling neural dynamics that may reflect activity across the broader therapeutic circuit rather than a single adjacent gray-matter structure.

In our previous work, we discovered a neural biomarker of response to VC/VS DBS in a cohort of 12 OCD patients.^20^ The biomarker quantified the temporal predictability of low-frequency activity recorded from the DBS target region in the background of everyday activities and demonstrated that disruption of pathologically predictable neural dynamics was associated with clinical response. These findings established proof of concept that chronic neural sensing could identify clinically meaningful biomarkers of DBS response. In this study, we extended neural predictability to a substantially larger longitudinal cohort and asked whether chronic neural sensing could provide clinically relevant biomarkers across multiple stages of DBS therapy. Specifically, we tested whether baseline neural dynamics before stimulation contain information about eventual treatment response, whether early changes in neural predictability following DBS activation provide evidence of therapeutic engagement before symptom improvement becomes apparent, and whether neural predictability can longitudinally distinguish clinical response states during chronic therapy. Together, these analyses establish a framework for neural biomarkers spanning the therapeutic arc of DBS, from pre-treatment forecasting to early target engagement and long-term treatment monitoring.

## Results

We analyzed neural data from 24 patients who underwent VC/VS DBS for treatment-resistant OCD (Fig. 1a; Supp. Fig. 1, 2; Supp. Table 1). The resulting dataset consisted of nearly 120,000 patient-hours of chronic neural recordings. Of the 24 patients, 14 achieved clinical response with DBS according to the Yale–Brown Obsessive Compulsive Scale (Y-BOCS; clinical response defined as Y-BOCS reduction ≥ 35%), four achieved partial response (Y-BOCS reduction ≥25% and <35%), and three did not achieve significant clinical improvement (Extended Data Table 1). Three additional patients were implanted within the past year and had not yet reached sufficient clinical follow-up duration to determine treatment outcome.

**Fig. 1:**
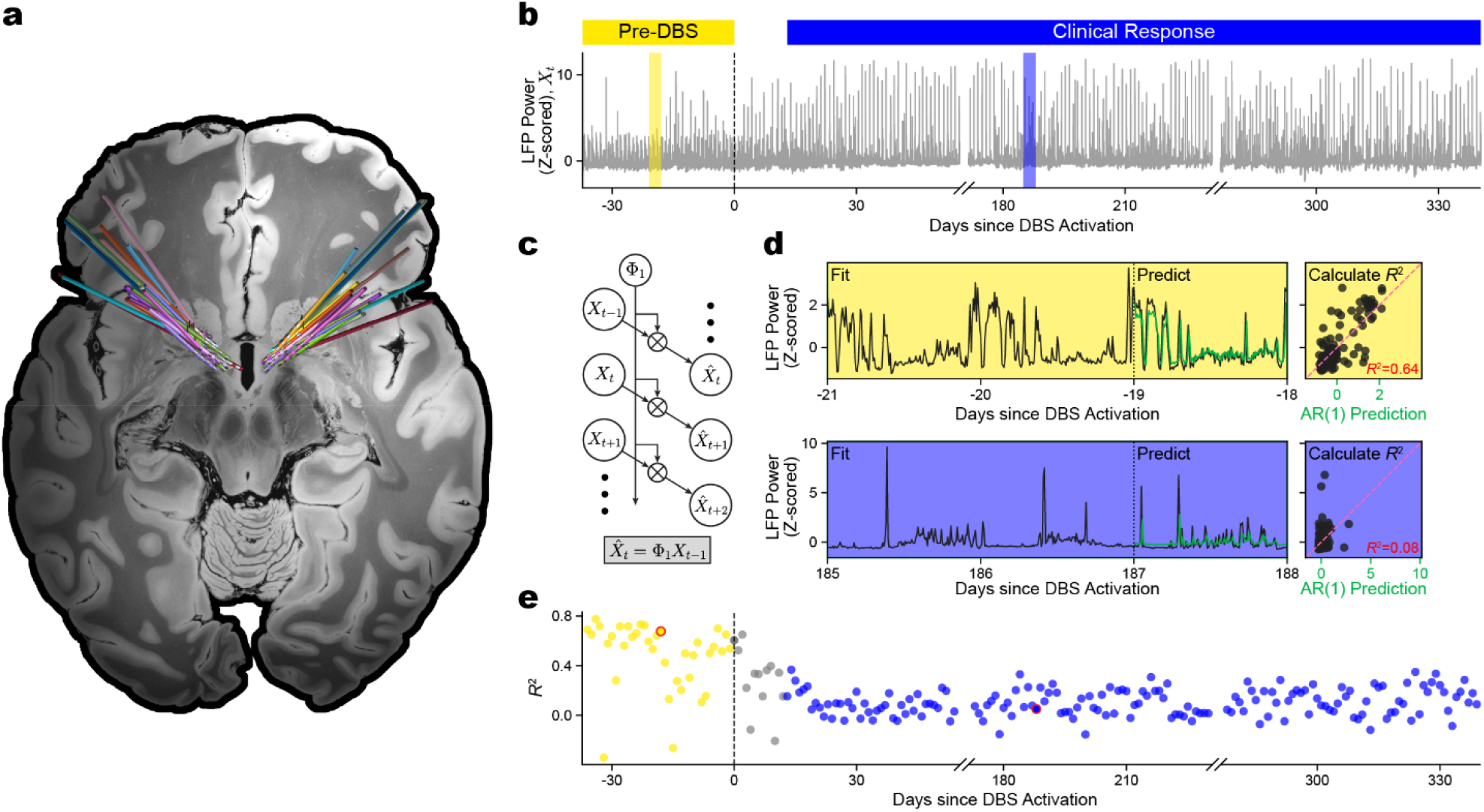
Prospective computation of neural predictability from chronic DBS recordings. **a**, Deep brain stimulation electrode locations for 24 patients with treatment-resistant obsessive-compulsive disorder (OCD) implanted bilaterally in the DBS target region. Lead trajectories are shown in Montreal Neurological Institute (MNI) space and color-coded by patient. **b**, Example longitudinal time series of daily z-scored 9 Hz local field potential (LFP) power for patient B011, spanning the pre-DBS period (yellow) through stable clinical response (blue). Highlighted intervals correspond to the examples shown in **d. c**, Schematic of the lag-1 linear autoregressive model used to predict each sample from the immediately preceding sample. **d**, Neural predictability was calculated within a three-day sliding window. The model was fit using the first two days and applied prospectively to predict the third day. The observed neural signals are shown in black, and the model predictions are shown in green. Neural predictability was quantified by computing the coefficient of determination (*R*^2^) between predicted and observed activity. Example windows from the pre-DBS period and stable clinical response show higher and lower agreement, respectively, between predicted and observed neural activity. **e**, Daily neural predictability across the full recording period for patient B011.

To quantify neural predictability, we developed a simple implementation that can be used to compute the biomarker prospectively from incoming neural recordings. We implemented a single autoregressive model architecture prospectively over the entire recording duration that predicted future neural activity using only preceding neural recordings, enabling prospective daily computation of neural predictability throughout DBS therapy. We used the Medtronic Percept DBS device to record average LFP power every 10 minutes within the 8.79 ± 2.5 Hz band (referred to hereafter as 9 Hz). Symptomatic periods were characterized by smooth, highly circadian fluctuations in 9 Hz power, whereas clinical response exhibited more frequent and transient high amplitude deflections (Fig. 1b). To quantify these differences in temporal organization, we applied a causal linear autoregressive model with a single lag term within a three-day sliding window (Fig. 1c). The model was trained using the preceding two days of neural activity and then used to predict the third day, yielding a measure of daily neural predictability (*R*^2^): the coefficient of determination between predicted and observed LFP activity (Fig. 1d, e).

### Neural predictability generalizes as a biomarker of DBS response

The neural predictability feature reliably distinguished responders from non-responders following VC/VS DBS in our cohort of 24 patients. Responders exhibited a significant reduction in neural predictability from the pre-DBS period to stable clinical response (*p* = 9.20 × 10^−19^), whereas non-responders did not show a significant change (*p* = 0.424; Fig. 2a, b; Extended Data Table 2, 3; Supp. Table 2). These findings extended our previous results in a substantially larger cohort using the simplified prospective implementation. Because baseline neural predictability varied substantially across patients (*µ* = 0.490; *α* = 0.154; Fig. 2c), we subtracted each patient’s pre-DBS baseline from the *R*^2^ time series. This change-from-baseline measure (Δ*R*^2^) reliably distinguished responders from non-responders (*p* = 2.62 × 10^−3^; Fig. 2d). Discriminability was specific to the left hemisphere (right hemisphere data did not distinguish response status; *p* = 0.600; Extended Data Fig. 1; Supp. Table 3, 4), reflecting hemispheric differences also described in previous studies.^34–37^

**Fig. 2:**
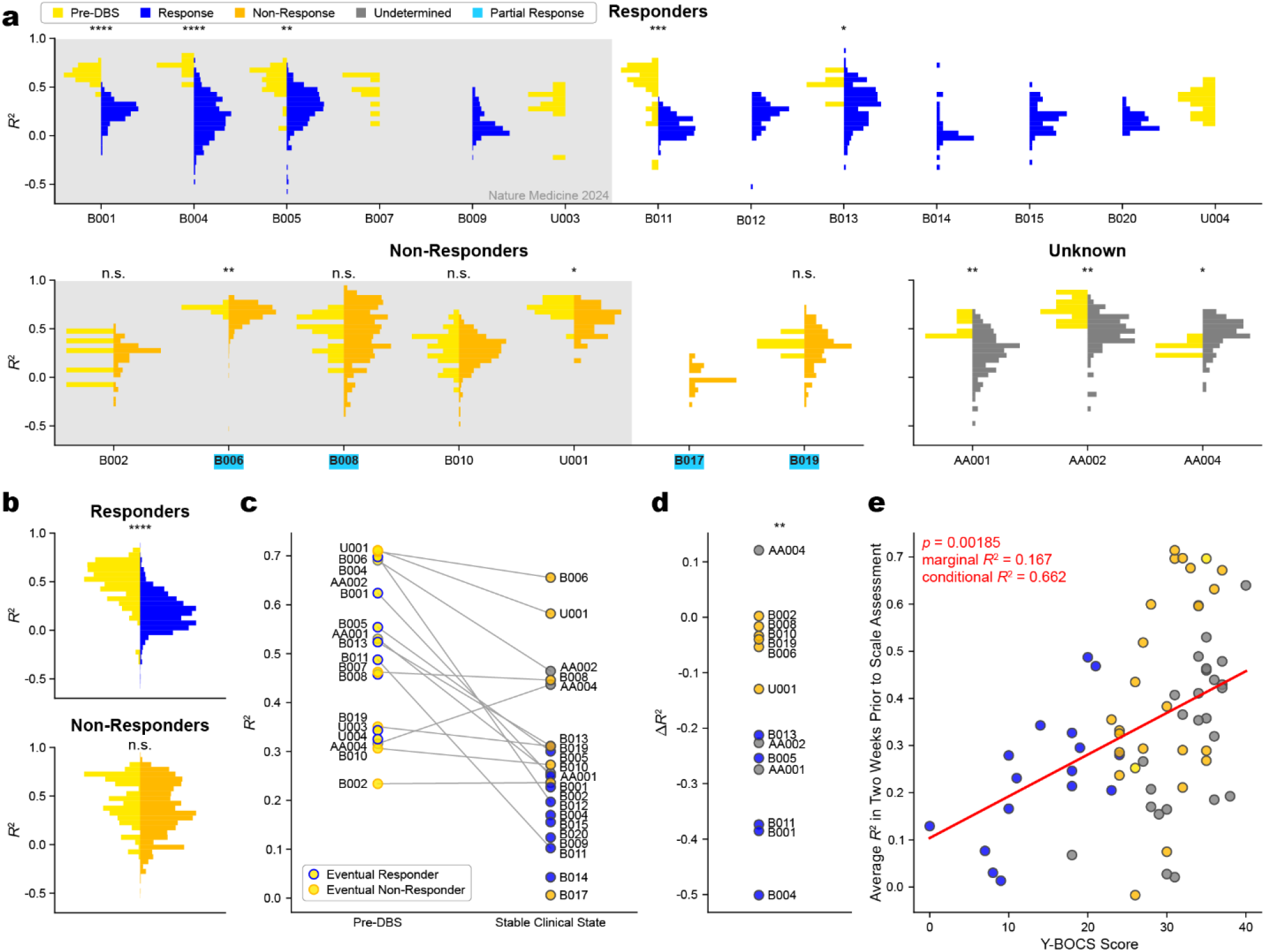
Neural predictability (*R*^2^) decreases with clinical response and relates to OCD symptom severity. **a**, Distributions of daily left hemisphere neural predictability during the pre-DBS and stable clinical states are displayed for individual patients. Patients included in the previous cohort20 are shown on a gray background. Pre-DBS values are shown in yellow, stable response values in blue, non-response values in orange, and values from recently implanted patients with undetermined outcomes in gray (N=3). Partial responder patient IDs are highlighted in light blue. Asterisks correspond *p* to values computed via Welch’s t-test with sample size correction. Statistical comparisons for individual patients are reported in Extended Data Table 2 and Supp. Table 2 for the left hemisphere and Supp. Table 3 for the right hemisphere. **b**, Pooled distributions of daily neural predictability (*R*^2^) for responders (N=13 with adequate data to calculate *R*^2^; N=14 total) and non-responders (N=7) during the pre-DBS and stable clinical state. Statistical results are reported in Extended Data Table 3 for the left hemisphere and Supp. Table 4 for the right hemisphere. **c**, The clothesline plot displays the average *R*^2^ during the pre-DBS and stable clinical states for all patients. **d**, The difference in average *R*^2^ of the pre-DBS and stable state shows a significant (*p* = 2.62 x 10^−3^) separation between responders and non-responders. **e**, Each Y-BOCS score (n=67) was plotted against the average *R*^2^ of the two weeks preceding the scale administration. Y-BOCS scores were significantly and positively associated with average preceding *R*^2^ values across patients.

We next evaluated whether neural predictability indexed symptom severity. Across patients, neural predictability was significantly correlated with Y-BOCS scores, with higher predictability associated with greater symptom severity via a mixed effects model (marginal *R*^2^ = 0.167; conditional *R*^2^ = 0.662; *p* = 1.85 × 10^−3^; Fig. 2e). During the transition period, one patient (B001) experienced a substantial temporary worsening of OCD symptoms despite persistently reduced neural predictability (Extended Data Fig. 2a, b). Because this episode occurred before the patient met criteria for stable response, these data were classified as transition and excluded from the binary classifier. A sustained relapse was also captured in B014 years after stable response had been established. Despite worsening symptoms during relapse, neural predictability remained low (Extended Data Fig. 2c, d). Together, these within-patient observations suggest that neural predictability may reflect a more slowly evolving physiological response to DBS rather than acute fluctuations in symptom severity.

### Changes in neural predictability classify clinical status

To evaluate whether neural predictability could classify stable clinical response vs. non-response on a daily basis (see Methods for determination of clinical status), we trained logistic regression models using leave-one-patient-out cross-validation. Separate models were trained using absolute neural predictability (*R*^2^) and the change-from-baseline feature (Δ*R*^2^). Because daily neural predictability exhibited short-term variability, we evaluated moving average windows ranging from 1 to 28 days. For the *R*^2^ feature, increasing window sizes generally slightly increased sensitivity and specificity, ultimately improving overall classification performance (Fig. 3a, b; Extended Data Table 4). Using a 14-day moving average, the classifier achieved an area under the receiver operating characteristic curve (AUROC) of 0.83 compared with 0.79 using single-day values (Fig. 3c, d; Extended Data Table 4). Performance was significantly better than chance classification (*p* < 1.00 × 10^−4^ for all moving average window sizes tested).

**Fig. 3:**
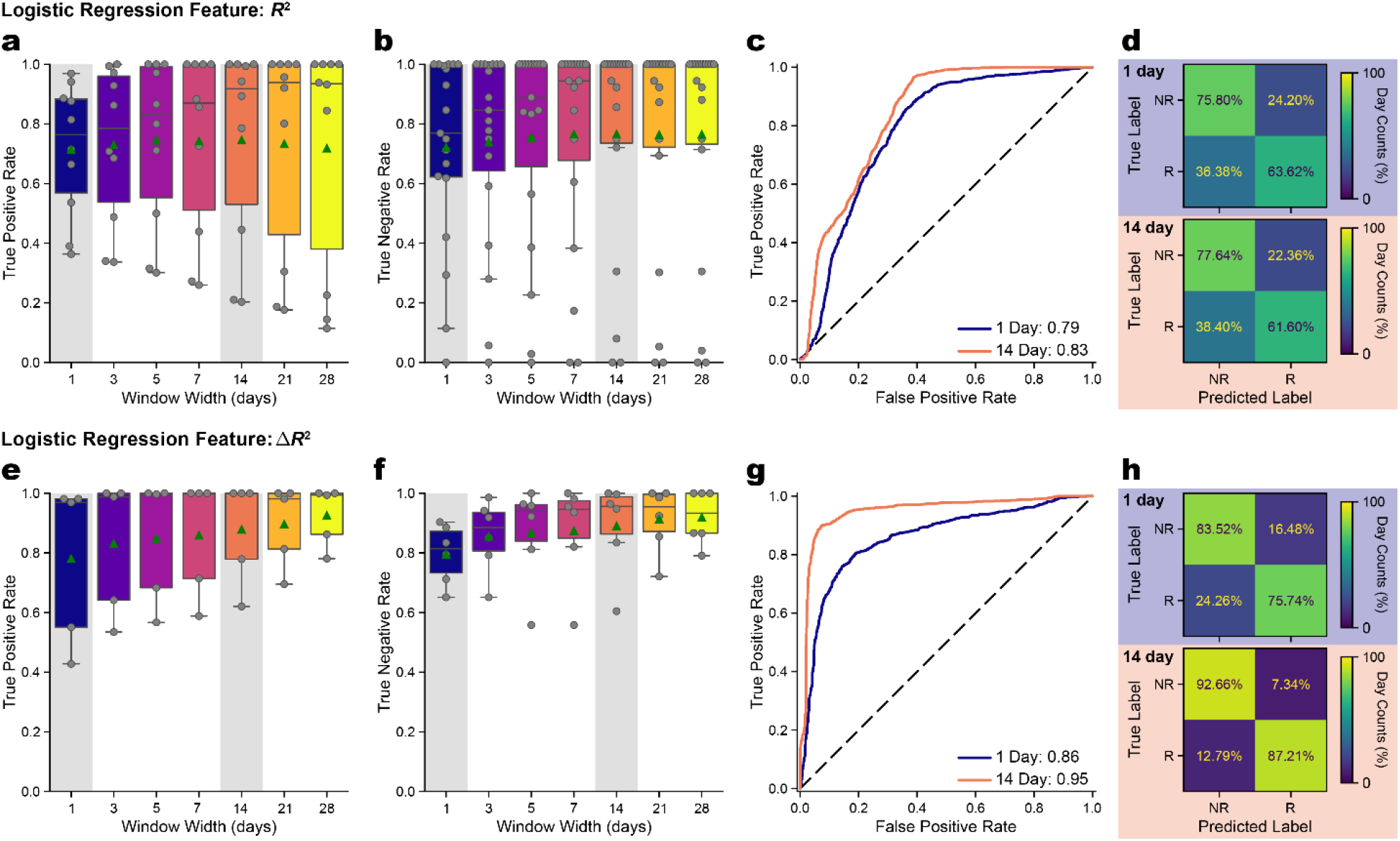
Neural predictability predicts clinical status with leave-one-patient-out cross-validation. We fit a logistic regression model on daily *R*^2^ and Δ*R*^2^ values to predict the clinical state associated with each day. To limit the effects of noise within the neural predictability features, we also applied moving averages across the biomarker signals with several widths. Operating points were computed for each feature and window width within each cross-validation fold such that the cutoff applied to the held-out patient was the one that equalized mean true positive and true negative rates across the training set patients. **a**, Box plots of the true positive rate (N=10) and **b**, true negative rate (N=17) for the logistic regression model using various moving average window sizes (1, 3, 5, 7, 14, 21, and 28 days) for *R*^2^ are shown. **c**, receiver operating characteristic (ROC) curves display the model performance of the daily and averaged (with a 14-day moving average window) *R*^2^ feature. **d**, Confusion matrices for the daily and averaged *R*^2^ models. The confusion matrices display the percentage of days predicted within each state. **e, f, g, h**, Analogous plots are shown with Δ*R*^2^ as the input feature for the logistic regression. For Δ*R*^2^, TPR N=5, TNR N=6. R, responders; NR, non-responders.

Subtracting each patient’s pre-DBS baseline neural predictability from the *R*^2^ time series substantially improved classification performance. Because Δ*R*^2^ requires pre-DBS recordings, 11 of the 24 patients were included in this analysis. Similar to the classification performance improvements we observed after smoothing the *R*^2^ feature with a moving average, classification performance with Δ*R*^2^ likewise improved after increasing the window size of the moving average (Fig. 3e, f). Again, using a 14-day moving average, we achieved an AUROC of 0.950 (Fig. 3g, h, Extended Data Table 4). Classifier performance was significantly greater than chance (*p* < 1.00 × 10^α4^ for all moving average window sizes tested, Extended Data Table 5). Because classification was performed at the level of patient-days, with adjacent observations highly autocorrelated within individuals and recording duration varying across patients, we additionally computed patient-weighted performance metrics so that each patient contributed equally. Patient-weighted performance remained high for all window widths in both the *R*^2^ and Δ*R*^2^ models. For the 14-day Δ*R*^2^ model, patient-weighted AUROC was 0.950, and balanced accuracy was 0.886 (Extended Data Table 4). These findings demonstrate that changes in neural predictability relative to each patient’s baseline provide substantially greater discriminatory power than absolute neural predictability. Performance remained comparable to more complex models, which did not result in meaningful improvements in sensitivity, specificity, or AUROC compared to the simpler single lag model formulation (Extended Data Fig. 3, Supp. Table 5).

### Early neural predictability changes forecast therapeutic response months before symptom improvement

To determine whether changes in neural predictability preceded clinical improvement, we analyzed the temporal trajectory of Δ*R*^2^ following DBS activation (Fig. 4a, b). In all five patients with recordings that spanned the pre-DBS period through clinical response, Δ*R*^2^ decreased below the logistic regression decision threshold within the first two weeks of DBS (Patient-fold regression thresholds reported in Extended Data Table 6). We found no significant difference in per-fold raw accuracies across the VC/VS versus VS/BNST implant trajectories (*p* = 0.923; Supp. Fig. 3). One additional patient (U003) also achieved clinical response but was initially programmed on an ineffective DBS contact.^38^ However, neural recordings were not available after the therapeutic stimulation contact was activated due to lack of sensing-compatibility.

**Fig. 4:**
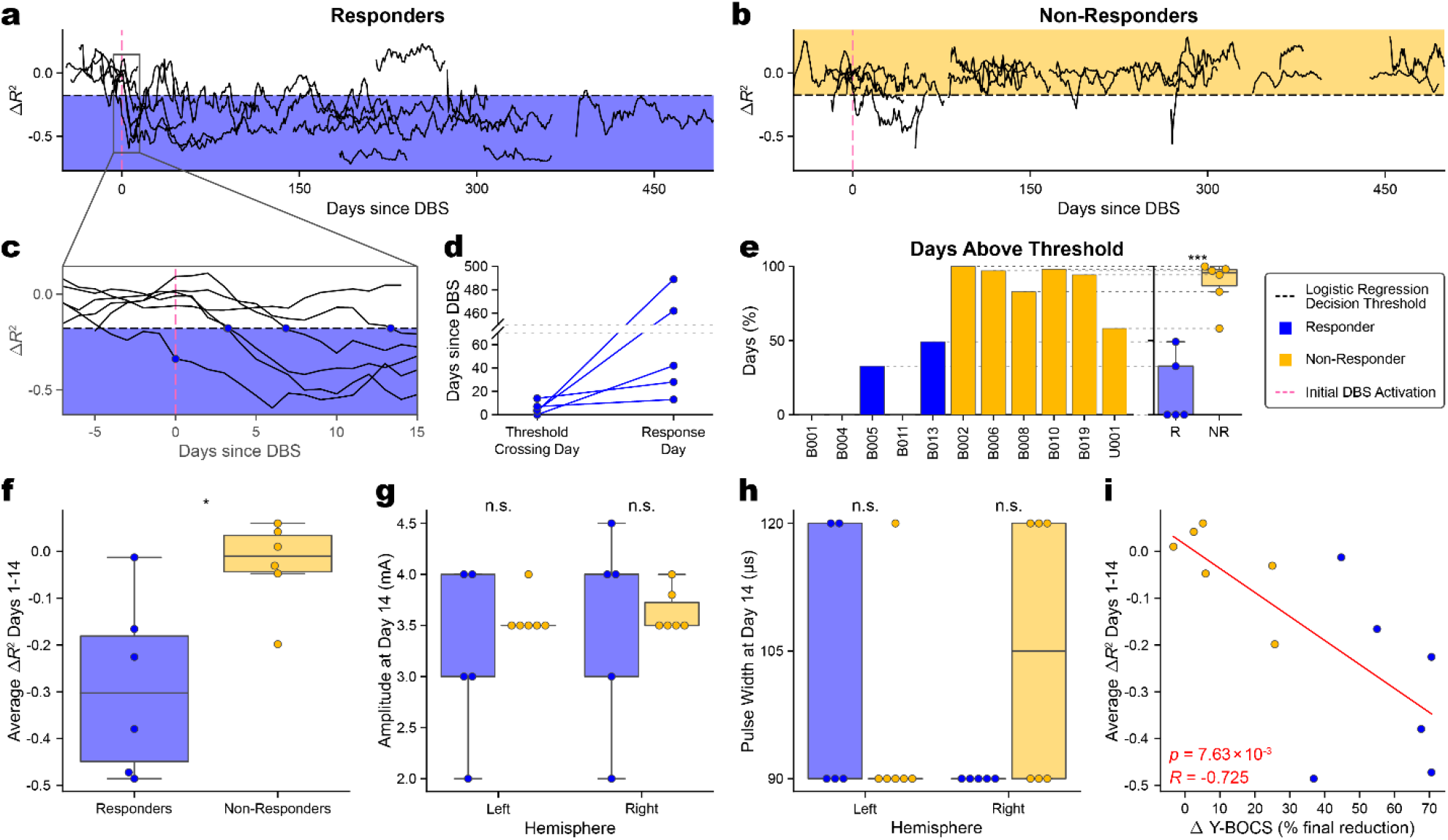
Changes in neural predictability precede symptom improvement. **a**, The 7-day moving average of *R*^2^ values over time are displayed for individual patients and time-aligned to DBS activation (vertical pink dashed line) for responders and **b**, non-responders. Decision boundaries were determined on a per-patient basis using leave-one-patient-out cross-validation. For visualization, the average logistic regression decision threshold across all folds (separately for responders and non-responders) is denoted by the horizontal dashed lines. **c**, Individual 7-day moving average Δ*R*^2^ trajectories crossed the logistic regression decision threshold for all five responders within 14 days, whereas non-responders rarely crossed the threshold. The day when the Δ*R*^2^ feature crossed the decision boundary is displayed for responders. **d**, A clothesline plot shows the day (since DBS activation) that the 7-day averaged Δ*R*^2^ crossed the patient-specific decision threshold, compared to the day each patient achieved stable clinical response (N=5 patients with both pre-DBS and post-response data). **e**, A bar plot and box plot show the percentage of Δ*R*^2^ values above the per-patient decision threshold after responder (N=5) or non-responder (N=6) determination (responders vs. non-responders: *p* = 6.33 × 10^−4^ via Welch’s two-sample two-tailed t-test). **f**, Box plots of the average Δ*R*^2^ in the two weeks after DBS activation show a significant difference between responders and non-responders (*p* = 1.70 × 10^−2^; Supp. Table 6). **g**, Box plots show DBS amplitude (*p* = 0.608 left, *p* = 0.783 right) and **h**, pulse width (*p* = 0.456 left, *p* = 0.302 right; Supp. Table 7) settings 14 days after DBS activation for both left and right hemispheres show no significant differences in stimulation parameters between responders and non-responders. Significance was determined using two sample, two-tailed Welch’s t-tests. **i**, Average Δ*R*^2^ values over the first two weeks after DBS activation are plotted against the overall Y-BOCS reduction recorded at most recent follow-up for each patient (see Extended Data Table 1 for follow-up durations). We performed linear regression to assess the association between Δ*R*^2^ after two weeks of DBS and overall Y-BOCS reduction (*p* = 7.63 × 10^−3^; *R* = − 0.725).

Although reductions in neural predictability occurred within the first two weeks following DBS activation (Fig. 4c), clinical improvement emerged much more gradually, with responders requiring an average of nearly 7 months to achieve stable response (*µ* = 206, *σ* = 220 days to response; Fig. 4d). Despite this delayed improvement in symptom severity, the average Δ*R*^2^ during the first 14 days after DBS activation was significantly lower in eventual responders than non-responders (Fig. 4f, Supp. Table 6). Responders also remained below the logistic regression decision threshold for a substantially greater proportion of the post-DBS period (responders: *µ* = 83.6%, *σ* = 20.7%; non-responders: *µ* = 11.6%, *σ* = 14.6% below the threshold; Fig. 4e), and separation between responder and non-responder trajectories became significant by day 4 after DBS activation (*p* = 0.0315; Supp. Video 1). These early differences were not explained by initial stimulation amplitude or pulse width, as these parameters did not differ significantly between groups (amplitude *p* = 0.608 left, 0.783 right; pulse width = 0.456 left, 0.302 right via Welch’s two-sample two-tailed t-test with correction for multiple comparisons; Fig. 4g, h, Supp. Table 7). Importantly, the average Δ*R*^2^ during the first 14 days after DBS activation was significantly correlated with each patient’s eventual reduction in Y-BOCS score at last follow-up (*R* = − 0.725; *p* = 7.63 × 10^α3^; Fig. 4i), demonstrating that early neural changes may forecast long-term therapeutic benefit months before clinical improvement.

### Baseline and DBS-induced circadian dynamics may distinguish DBS responders

Because our neural predictability feature does not fully capture daily temporal dynamics, we next investigated the impact of DBS on the circadian organization of neural activity. Responders exhibited marked changes in circadian neural architecture following successful DBS, whereas non-responders showed relatively stable patterns (example patients shown in Fig. 5a). Spectral analysis revealed significant reductions in both the 12-hour and 24-hour periodic components following clinical response (12-hour: *p* = 0.0371, 24-hour: *p* = 0.0371), while these circadian rhythms remained largely unchanged after DBS in non-responders (12-hour: *p* = 0.375, 24-hour: *p* = 0.375; Fig. 5b, c, Supp. Table 8). These findings indicate that effective DBS is associated with substantial reorganization of VC/VS circadian dynamics.

**Fig. 5:**
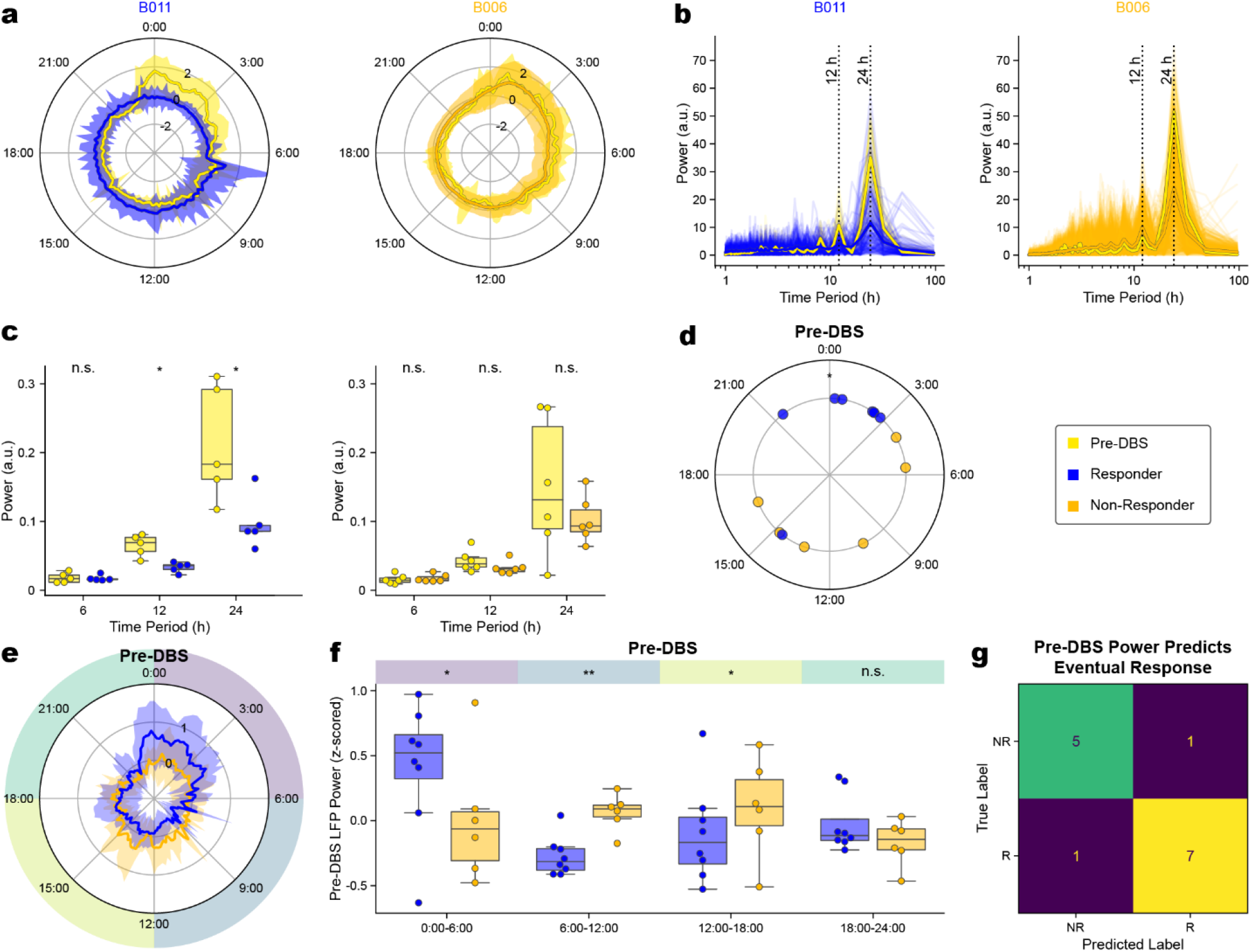
Effective DBS reorganizes 24-hour LFP dynamics and reveals baseline features associated with treatment response. **a**, Example 24-hour profiles of 9 Hz power (mean and standard deviation for each time bin) for an example responder (B011) and non-responder (B006) across the pre-DBS and stable clinical states are plotted on polar axes. **b**, Corresponding Welch power spectral density (PSD) estimates showing power at periodicities spanning the 24-hour cycle. **c**, Distributions of PSD at the 6-hour, 12-hour, and 24-hour components before DBS and in the stable clinical state for responders (N=5) and non-responders (N=6) with neural recordings both before DBS and after clinical state determination. Pre-versus post-DBS comparisons were performed using paired two-tailed t-tests with correction for multiple comparisons. Adjusted *p* values for 6-hour, 12-hour, and 24-hour components were *p* = 0.776,0.0371,0.0371 for responders and *p* = 0.375,0.375,0.375 for non-responders. **d**, Mean left-hemisphere pre-DBS acrophase for each patient is shown. Baseline acrophase differed significantly between eventual responders and non-responders (*p* = 0.0211; two-sample Mardia–Watson–Wheeler test). **e**, Mean pre-DBS 9 Hz power across the 24-hour cycle for eventual responders and non-responders, averaged within each 10-minute time bin. **f**, Pre-DBS power values averaged across four 6-hour time bins for eventual responders (N=8) and non-responders (N=6) with available pre-DBS neural data. Group differences within each time bin were assessed using a mixed linear model with correction for multiple comparisons; adjusted values for [0:00-6:00], [6:00-12:00], [12:00-18:00], [18:00-24:00] were *p* = 0.0213, 6.82 × 10^−3^, 0.0213, 0.269, respectively. **g**, Confusion matrix for prediction of eventual responder status using pre-DBS power values as inputs to a linear discriminant analysis model with leave-one-patient-out cross-validation. Within each fold, time bins associated with responder status were identified using only the training patients and subsequently used to classify the held-out patient.

Finally, we explored differences in baseline neural activity between eventual responders and non-responders to determine whether features of underlying neurophysiology before DBS activation are related to treatment outcome. Remarkably, circadian neural organization differed between eventual responders and non-responders even before DBS therapy began. Eventual responders exhibited significantly different baseline left-hemisphere neural acrophases relative to non-responders (*p* = 0.0211; Fig. 5d, Supp. Table 9). We additionally observed distinct time-of-day patterns of LFP power in the early morning (0:00-6:00; *p* = 0.0213), late morning (6:00-12:00; *p* = 6.82 × 10^α3^), and afternoon (12:00-18:00; *p* = 0.0213) (Fig. 5e, f, Supp. Table 10). Neural power in the evening (18:00-24:00) was not significantly different between responders and non-responders (*p* = 0.269). We then trained a leave-one-patient-out linear discriminant analysis (LDA) classifier using baseline power from the 6-hour time-of-day bins that differed between responders and non-responders within each training fold. The model accurately classified eventual responder status in all but two of the 14 patients, significantly greater than chance (AUROC=0.833, *p* = 0.00980; Fig. 5g). These findings suggest that circadian organization of neural activity prior to stimulation may contain clinically relevant information about a patient’s likelihood of responding to DBS therapy.

## Discussion

A major challenge in psychiatric neuromodulation is the identification of neural biomarkers that can guide clinical decision-making across the full course of therapy. Here, we identify complementary neural signatures at three clinically distinct stages of DBS treatment. Before stimulation, baseline circadian neural dynamics distinguished eventual responders from non-responders, demonstrating that pre-treatment physiology contains information relevant to treatment outcome. During the first two weeks after DBS activation, reductions in neural predictability forecast eventual therapeutic benefit months before clinical improvement became apparent, providing an early physiological marker of target engagement. During chronic therapy, neural predictability distinguished stable response from non-response and tracked longitudinal clinical state. Together, these findings support neural predictability as a candidate biomarker of DBS target engagement and clinical outcome that motivates a broader framework in which successful DBS may restore flexibility to pathological neural dynamics within cortical and subcortical reward and motivational circuits.^31,39^

Interpreting the biological basis of neural predictability is critical for understanding its relationship to OCD pathophysiology. Unlike many proposed DBS biomarkers that quantify the momentary magnitude of neural power within a specific frequency band, neural predictability quantifies the temporal organization of neural activity, capturing how neural dynamics evolve over time rather than the moment-to-moment amplitude of activity. Neural predictability is highest when DBS target activity exhibits highly stereotyped temporal dynamics and can therefore be viewed as an interpretable measure of temporal neural flexibility, with lower predictability reflecting less stereotyped temporal dynamics. Successful DBS was consistently associated with reduced neural predictability, suggesting a transition from rigid, repetitive neural dynamics toward more flexible patterns of activity, consistent with prevailing models of obsessive-compulsive disorder that emphasize pathological inflexibility within circuits responsible for governing fear response, reward sensitivity, motivation, and controlled decision-making.^39,40^ Rather than reflecting disorganized neural activity, the emergence of greater temporal variability may indicate restoration of the dynamic neural repertoire required for adaptive cognition and behavior. We therefore propose that neural flexibility represents one potential mechanism linking DBS target physiology to the approach-avoidance framework.^41^ Specifically, we hypothesize that restoration of flexible neural dynamics enables patients to disengage from compulsive avoidance and re-engage with motivationally salient environmental stimuli. Future studies integrating chronic neural recordings with dense behavioral phenotyping will be required to directly test this mechanistic hypothesis.

Our findings support the view that neural variability can represent an adaptive property of healthy brain dynamics rather than simply noise. Neural systems must balance stability with flexibility: overly stereotyped activity may constrain transitions among functional states (e.g., as in coma^42^), whereas excessive variability may impair reliable information processing (e.g., as in devastating epilepsies^43^ and neurodegenerative disorders^44^). Across scales, variability and heterogeneity can expand the repertoire of accessible neural states and support flexible responses to changing internal and environmental demands. From this perspective, the reduction in neural predictability observed following effective DBS may reflect a shift away from an overly constrained dynamical regime toward a more flexible state, rather than a nonspecific increase in neural disorder. Such a homeostatic state could preserve sufficient stability for reliable information processing while allowing the system to explore a broader range of neural configurations. This framework provides a biologically plausible interpretation of reduced neural predictability as a marker of restored dynamical flexibility and may help explain why successful DBS is associated with less stereotyped neural activity.

Linking neural dynamics with clinical symptoms remains a fundamental challenge in the development of psychiatric biomarkers,^45,46^ in part due to the historical confinement of electrophysiology studies to laboratory or clinical settings where longitudinal, naturalistic symptom dynamics are difficult to capture.^7^ Our findings suggest that neural predictability contains information relevant both for long-term treatment response and overall severity of symptoms. However, establishing whether it can track fine-grained symptom fluctuations will require behavioral measurements with substantially higher temporal resolution. A fundamental limitation is the temporal resolution mismatch between continuous neural sensing and conventional clinical assessments. Although neural activity is sampled every 10 minutes, behavioral measurements such as the Y-BOCS are typically obtained weeks to months apart,^47^ limiting the ability to resolve dynamic relationships between neural activity and symptom fluctuations. Furthermore, clinical improvement following DBS often evolves gradually and nonlinearly,^26,27,48^ further obscuring the temporal relationship between neural physiology and behavior. Consistent with this interpretation, despite worsening clinical symptoms during the single relapse period captured in our cohort in B014, as well as during the significant worsening in severity observed before stable response in B001, neural predictability remained relatively stable. Although definitive conclusions cannot be drawn from these limited observations, these examples support the possibility that neural predictability primarily reflects sustained target engagement or longer-term circuit reorganization rather than moment-to-moment symptom expression. Distinguishing these possibilities will require larger longitudinal cohorts with more frequent behavioral assessments and additional instances of rapid symptom changes captured during chronic neural sensing. Integrating chronic neural sensing with high-resolution behavioral measurements, including wearable devices, ecological momentary assessments, and passive smartphone-based monitoring, may enable a more complete understanding of how neural dynamics relate to symptom expression and ultimately support more personalized and physiology-guided neuromodulatory therapies.^49^

Determining whether neural predictability reflects successful target engagement has important implications for DBS programming. Although successful therapy was consistently associated with reductions in neural predictability, establishing a causal relationship will require prospective studies that evaluate how this biomarker changes following systematic adjustments in stimulation parameters, contact selection, and lead location. Demonstrating that neural predictability reliably responds to these programming interventions would provide strong evidence that it reflects target engagement and could establish an objective physiological endpoint to guide DBS programming. Ultimately, chronic biomarkers such as neural predictability may complement acute physiological biomarkers measured during implantation^50–53^ or early programming.^54^ These complementary biomarkers may support different clinical decisions at different stages of therapy: baseline physiology may help identify patients most likely to benefit, early changes in neural predictability may provide evidence that stimulation is engaging therapeutic circuitry, and chronic sensing may provide an objective measure of longitudinal treatment state.

The anatomical location of these recordings also informs their interpretation. Although DBS in this region is conventionally described using gray matter landmarks such as the VS or BNST, the chronic sensing contacts in this cohort were centered within, or were just adjacent to, the VC white matter. Some overlapped with gray matter of the VS, BNST, globus pallidus, and ventral pallidum, as well as white matter of the anterior commissure. Moreover, the relationship between reduction in neural predictability and clinical outcome was observed across trajectories targeting the VC/VS and VS/BNST, despite differences in the subjacent gray matter structures. These findings therefore suggest that this neural biomarker captures physiological dynamics within a distributed circuit, which includes nearby gray (VS, BNST, globus pallidus externus, globus pallidus internus, ventral pallidum) and white matter (VC, anterior commissure) components (Fig. 1a; Supp. Fig. 1, 2; Supp. Table 1), rather than exclusively reflecting activity from a single neighboring nucleus.

The physiological processes underlying these distributed changes remain incompletely understood, particularly because the narrow-band recordings used here do not capture the full bandwidth of neural activity. Our analyses suggest that reductions in neural predictability appear to align with the emergence of transient high amplitude deflections in DBS target activity which disrupt otherwise highly stereotyped circadian patterns. These deflections also coincide with the circadian reorganization observed in responders, including shifts in acrophase and reduced nocturnal neural power, suggesting that they may represent a broader reorganization of VC/VS circuit dynamics following effective DBS. These naturalistic 24-hour patterns may reflect both endogenous circadian processes and behaviorally structured daily activity. We hypothesize that these transient events reflect engagement of reward- and motivation-related processes within distributed circuits traversing the ventral capsule, including cortical, basal ganglia, and extended amygdala networks involved in reward processing and motivated behavior.^55–57^ As patients recover from the rigid, avoidance-dominated behavioral state characteristic of severe OCD,^58–60^ increased engagement with motivationally salient aspects of their environment may give rise to these transient neural events and the associated reduction in neural predictability.

Several limitations should be considered when interpreting these findings. First, although this represents a relatively large longitudinal sensing cohort for psychiatric DBS, the number of patients remains modest. Recording availability was also heterogeneous across participants, with only a subset contributing pre-DBS data or recordings spanning both pre- and post-treatment states. However, this heterogeneity in data availability is a limitation that is inherent to any real-world study that is not part of a rigorously controlled trial. Second, clinical state was defined using intermittently sampled Y-BOCS assessments, limiting temporal precision in relating neural dynamics to symptom fluctuations. This intermittency is also a reflection of the real-world nature of conducting clinical assessments outside of a controlled trial, as patients are generally averse to frequent completion of burdensome assessments. Finally, BrainSense Timeline recordings were restricted to a narrow frequency band in a single region and therefore do not capture the full spectral or circuit-level neurophysiology underlying changes in neural predictability. Prospective validation in independent, larger cohorts with denser behavioral sampling and full bandwidth neural recordings will be necessary to determine the generalizability and clinical utility of these biomarkers.

Collectively, these findings establish a framework for neural signatures spanning the therapeutic arc of DBS: pre-treatment neural features that forecast eventual response, early neural changes that index therapeutic engagement, and chronic biomarkers that track clinical state during ongoing therapy. Although developed in OCD, this conceptual framework may extend to other psychiatric disorders treated with sensing-enabled neuromodulation. Realizing this vision will require prospective validation in larger multi-center cohorts, integration of high-resolution behavioral phenotyping with chronic neural recordings, and continued advances in sensing-enabled neuromodulation technologies. As these capabilities mature, objective physiological biomarkers have the potential to fundamentally transform psychiatric neuromodulation from a predominantly empirical, symptom-driven practice toward a future in which objective neural biomarkers complement clinical judgment to guide individualized therapy delivery.

## Methods

### Study design

This study included 24 participants who underwent DBS treatment for OCD, 12 of whom were introduced in our previous work^20^ (B001-B010, U001-U003) and 12 of whom are newly reported here (B011-B020, AA001-AA004, and U004). All procedures were approved by the institutional review boards at Baylor College of Medicine (IRB H-48392; H-56119) and the University of Utah (IRB 00169174). Patients were not compensated for any research described here. All patients qualified for DBS for OCD under the same well-accepted criteria: diagnosed with severe OCD more than 5 years ago, failed at least 3 trials of SSRIs of appropriate dose and duration, failed clomipramine and antipsychotic augmentation, as well as at least one adequate course of expert exposure and response prevention (ERP) therapy. Patients B012, B014-B017, and B020 underwent battery replacement surgeries where their previous implantable pulse generator (IPG) devices were replaced with Medtronic Percept devices. As a result, we were only able to collect data after their battery replacements, about 2+ years after DBS activation. Three patients (AA001, AA002, and AA004) were implanted recently and have not had sufficient clinical follow-up (>1 year) to determine a clear clinical status. Patient demographics are included in Extended Data Table 1.

### DBS surgery

DBS leads (Medtronic 3387 [N=8] or SenSight [N=16]; Supp. Table 1) were placed bilaterally in the ventral capsule (VC) region. Targeting was performed directly using the pre-operative magnetic resonance imaging (MRI). DBS leads were implanted using one of two candidate trajectories through the VC. One trajectory positioned the distal lead tip near the ventral capsule/ventral striatum border, whereas the second positioned the tip near the ventral capsule/bed nucleus of the stria terminalis (BNST) border. The distance between these targets is only a few mm. Final trajectory selection was informed by awake intraoperative behavioral testing as previously described.^52^ Although the distal anatomical targets differed slightly between trajectories, the contacts used for chronic sensing in this study were located within or adjacent to the ventral capsule white matter, with the most ventral contacts variably approaching the ventral striatum or BNST region depending on the selected trajectory. The pair of leads was connected to extensions that were tunneled to a Medtronic Percept PC or Percept RC IPG.

### Imaging protocol and DBS electrode localization

Preoperative high-resolution T1-weighted magnetic resonance imaging (MRI) was obtained on a 3 Tesla Siemens MAGNETOM Prisma Fit scanner, and postoperative noncontrast helical computed tomography (CT) was acquired on the day of electrode implantation. Electrode localization was performed using Lead-DBS v3.0.^61^ Preoperative MRI and postoperative CT datasets were first linearly aligned with Advanced Normalization Tools (ANTs),^62^ then nonlinearly transformed into MNI 2009b standard space using the ANTs Symmetric Normalization (SyN) algorithm. To compensate for postoperative brain deformation, brain shift correction was applied using the coarse mask approach.^63^ Electrode trajectories were automatically reconstructed using the Precise and Convenient Electrode Reconstruction for Deep Brain Stimulation (PaCER) toolbox and subsequently reviewed and manually adjusted as needed.^64^ Finally, electrode locations across patients were simultaneously visualized using Lead Group in a 7 Tesla Ex vivo 100um Brain Atlas.^65^ Active electrode locations are shown in Supp. Fig. 1, 2 and Supp. Table 1.

### Determination of clinical status

Clinical status was determined by the Yale–Brown Obsessive Compulsive Scale (Y-BOCS). The scales were administered before surgery and periodically throughout the course of treatment under the supervision of E.A.S. A baseline Y-BOCS measurement was captured before DBS surgery, and clinical status was subsequently defined by the percent change from the baseline Y-BOCS. Between assessments, each Y-BOCS score was carried forward and treated as constant until the next assessment.

Stable responders were defined as patients who achieved ≥35% reduction in Y-BOCS from baseline and subsequently maintained at least a partial clinical response (≥25% reduction) for ≥12 months. Temporary fluctuations into the partial response range were permitted if they lasted <6 months and were followed by a return to the response range (≥35% reduction). Stable responder status was determined retrospectively based on the 12-month durability criterion. For patients who met this criterion, the stable response state was defined as beginning with the first Y-BOCS assessment demonstrating ≥35% reduction that was subsequently followed by ≥12 months of maintained clinical benefit as described above.

Non-responder patients who achieved partial response (Y-BOCS reduction ≥25% and <35%) at any point after DBS activation were annotated in Fig. 2. Non-responders were defined as patients who never achieved stable response criteria after receiving at least one year of DBS stimulation.

The period between DBS activation and the first qualifying response assessment was considered as a transition period during which ground truth clinical status was difficult to assign given the sparse nature of Y-BOCS assessments. Accordingly, we labeled clinical status as undetermined for the three patients in our cohort who had received less than one year of DBS therapy by the time we completed our analyses. The stable clinical state refers to the time after achieving stable response in responders and the entire post-DBS period for non-responders.

Based on the criteria above, all recorded days of neural data were labeled as either stable response (all recorded data after stable response was achieved) or non-response (all recorded data after DBS activation in non-responders). Stable response and non-response periods comprise the positive and negative classes in our binary classification analyses, respectively (Figs. 2, 3, and 5). Data from undetermined and transition periods were omitted from our classification analyses.

One patient (B014), who had previously met the criteria for a stable response, experienced a sustained relapse approximately 1.5 years after initially responding. This period lasted 292 days before the patient remitted, yielding 37 days of recorded neural data during the relapse. An additional 37 days of neural data were collected after the relapse period ended.

Because the relapse represented a sustained loss of clinical response, neural data recorded during this period were assigned to the non-response state, while the patient remained classified as a stable responder at the patient level. We did not capture neural data during sustained relapse for any other patients in our cohort.

For the patient-level clinical status classification analysis, data from patients with undetermined clinical status were excluded. Data collected after stable responders were included in the positive class, and data collected from non-responders (including partial responders) were included in the negative class.

### Device setup and data collection

The Percept devices were configured to record a continuous time series of average LFP power in units of microvolts peak within a user-specified 5 Hz frequency band every 10 minutes (144 per day), using the BrainSense Timeline modality. The recordings were performed with a bipolar sensing montage centered on the active stimulation contact(s). Therefore, recording was only possible when the stimulating contact was set as one or both of the middle contacts on the lead.

We configured the devices to record LFP power in the band centered at 8.79 Hz (referred to in the main text and hereafter as 9 Hz). The 10-minute power averages were stored onboard the device and downloaded upon connection to the Medtronic DBS Clinician Programmer Application, which was subsequently offloaded to our local PHI-compliant database.

The data must be manually downloaded by a member of our team or a Medtronic representative, so regular in-person visits were necessary to collect data. The Percept RC and PC IPGs have respective onboard storage limits of 35 and 60 days, after which the oldest recordings are overwritten. These storage limits resulted in occasional periods of lost data when patients could not regularly visit the clinic or meet a Medtronic representative.^66^

### Data pre-processing

BrainSense Timeline data, stimulation parameters, and associated metadata were extracted from the downloaded JSON files.

Timestamps were converted from UTC to the home time zone of each patient. Outliers resulting from overvoltage artifacts were identified and corrected using the principled overvoltage event removal method we described previously.^67^ One patient (B015) experienced a period of 37 days where overvoltage events dominated the time series, with over 20% of data on each day compromised by such artifacts. Because we could not confidently recover the true time series in this case with such a high proportion of outliers, we excluded these data from analyses. After overvoltage correction, small gaps in the data were characterized by missing timeline values for up to 12 consecutive samples and interpolated with a piecewise cubic Hermite spline (PCHIP interpolation). Larger gaps of more than two hours were not interpolated and left empty. Finally, because baseline LFP and stimulation amplitude generally increased over time for both responders and non-responders, LFP power values were z-scored within each day using daily mean and standard deviation values.

### Linear autoregressive model construction

We used a lag-1 linear autoregressive (LinAR-1) model to predict neural power at each 10-minute timepoint. A generic linear autoregressive model can be expressed as (1):

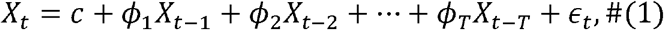

where *X*_*t*_ is the value of the time series at time *t, c* is the steady-state bias term learned by the model, *ϕ*_*k*_ are the coefficients for each of the *T* lag terms of the model, and *ϵ*_*t*_ is the residual error term representing the difference between the model’s predicted and actual values at time *t*. Our model used only *T* = 1 lag term, and we did not include a constant bias term because our data was z-scored and therefore had zero mean. Thus, the model can be expressed as (2):

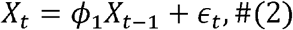

where *ϕ*_1_ is the only learnable parameter of the model, and *X* represents the pre-processed neural LFP power time series.

We trained the LinAR-1 model with a 3-day sliding window and 1-day stride on the LFP power time series. Within each window, the model’s parameter was learned by fitting an ordinary least squares regression model on the first two days of pre-processed LFP power values. Then, the model was applied to the data within the window’s final day. No predictions were made when the training set contained fewer than 144 samples (24 hours) or the test set contained fewer than 72 timepoints (12 hours) due to small sample sizes producing noisy and unstable predictions. One patient (U002) had extremely limited neural recordings (*µ* = 9.02, *σ* = 2.86 hours per day, n=15 days). Therefore, their data were excluded.

### Feature calculation

We calculated the daily coefficient of determination (*R*^2^) between the autoregressive model’s predicted time series values and the corresponding ground truth time series values of each day. The daily *R*^2^ score was calculated using the scikit-learn Python package and can be expressed as (3):

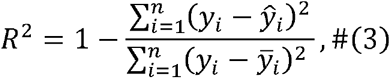

where *y*_*i*_ is the true LFP power value, *ŷ*_*i*_ is the predicted power value at sample *i* and 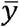 is the mean of the true values for a given day. Since the *R*^2^ values were calculated separately per day, *n* was maximally 144 on any given day. Due to the methodology of the *R*^2^ calculation, negative *R*^2^ values can arise if the daily sum of the squared residuals is greater than the total sum of squares around the observed mean, such that the model achieves worse performance than predicting the daily mean.

The learned parameter *ϕ*_1_ was generally slightly less than 1 in our cohort. Thus, the model’s prediction for the time series was a delayed and slightly attenuated version of the ground truth time series. The *R*^2^ score between the prediction and the ground truth value is therefore relatively large when two consecutive samples contain similar values and relatively small when the signal has a high rate of change.

We also subtracted each patient’s pre-DBS average *R*^2^ from the *R*^2^ time series to quantify the change in *R*^2^ each patient experienced from baseline. To do this, we took the average *R*^2^ of each patient’s pre-DBS state and subtracted it from each sample in the post-DBS *R*^2^ time series. This resulted in a new daily feature, termed Δ*R*^2^. This was only possible for patients with pre-DBS data available (N=11). Pre-DBS data were removed from logistic regression model runs using the Δ*R*^2^ feature to prevent leakage. This implementation differed from our previous study^20^ and explains the slight difference in model performance when using the Δ*R*^2^ feature with the same model on the previous dataset (Extended Data Table 6, Supp. Table 5).

To generate the video illustrating Δ*R*^2^ progression (Supp. Video 1), patient timelines were aligned to the day of DBS activation (day 0). The average Δ*R*^2^ for each patient was then computed daily, from day 0 through each subsequent day across the first 500 days after DBS activation. Welch’s t-tests were used to compare the Δ*R*^2^ distributions between eventual responders and non-responders on all 500 days and corrected using Benjamini-Hochberg FDR for multiple comparisons.

### Logistic regression clinical symptom state classification

We trained models to classify symptom-burdened and symptom-unburdened status each day. We applied a causal *k*-day moving average to the *R*^2^ and Δ*R*^2^ feature time series, with *k*∈ {1, 3,5, 7, 14,21, 28} to determine a suitable window length for classification, using a leave-one-patient-out cross-validation. We then compared regression model performance between the daily *R*^2^ and Δ*R*^2^ features to the moving average versions. The 14-day window was selected as a representative clinically interpretable timescale for subsequent analyses rather than as a formally optimized hyperparameter. We computed the receiver operating characteristic (ROC) curve, area under the ROC curve (AUROC), balanced accuracy, true positive and true negative rates, and a confusion matrix for each model’s combined predictions.

To convert predicted probabilities into binary classifications, we derived an operating point within each cross-validation fold. For a given feature and window width, the cutoff applied to a held-out patient was the equal error rate (EER) operating point for the out-of-fold predictions of the remaining patients. This EER which minimized the absolute difference between the mean true positive rate and mean true negative rate. Operating points were computed independently for each feature and window width. Because no patient’s own labels contributed to the cutoff applied to that patient, the decision boundary is derived entirely from training data.

Because patients contributed different numbers of recording days, and daily values within a patient are strongly autocorrelated, pooled day-level metrics can overstate the precision of classification performance. We therefore also computed patient-weighted performance metrics, in which each held-out day was weighted by the reciprocal of the number of held-out days contributed by that patient, so that every patient contributed equal total weight. Patient-weighted AUROC, balanced accuracy, true positive rate, and true negative rate were computed from the same leave-one-patient-out predictions and at the same held-out operating points as the day-level metrics (Extended Data Table 4).

Classifier performance was also compared to chance classification performance. We randomly shuffled state labels among the entire dataset using two methods: (1) a random shuffle across all patients and (2) via a random circular shift per patient. We only applied the circular shift to models using the daily *R*^2^ feature, as both the Δ*R*^2^ and averaged features were limited to a single class (i.e., either responder or non-responder). We then used a leave-one-patient-out cross-validation strategy on the shuffled data to evaluate the random performance. This was repeated 10,000 times to create a random classifier distribution. The AUROC, balanced accuracy, true positive, and true negative rates were calculated for each run to form chance distributions. The true classifier’s performance metrics were compared to the chance distributions and used to *p* calculate values, representing the percentage of chance distribution that was greater than the true performance. The resulting statistics are reported in Extended Data Table 5.

### Statistical methods

All inferential statistics were two-tailed unless otherwise noted, and statistical significance was assessed at *α* = 0.05. Independent two-sample comparisons (e.g., comparing responders versus non-responders on a single summary metric) were performed with Welch’s two-sample t-test. Within-subject comparisons (e.g., pre-versus post-DBS within the same subject) were performed with the paired t-test (Fig. 5c). For group/paired mean comparisons, we reported standardized effect sizes (Hedges’ G), *p*-values, and 95% confidence intervals to convey the magnitude and precision of each finding (Extended Data Tables 2, 3; Supp. Tables 2, 3, 4). To account for multiple comparisons, we applied the Benjamini–Hochberg procedure for controlling the false discovery rate (FDR)^68^ to each family of comparisons between responders and non-responders (Figs. 4g-h, 5c, and 5f).

Effective sample size correction (ESS) was applied to estimate the true amount of data within each time series. This uses autocorrelation to identify how much redundant information is contained within consecutive items of the time series (4):

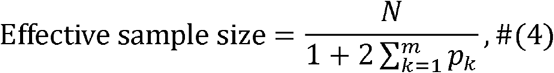

where *N* is the original sample size, and *p*_*k*_ is the autocorrelation at lag *k* up to a maximum lag of *m*. In our analysis, we chose a maximum lag of 30 days or the number of days present, whichever was lower. This correction technique was used for the within-patient comparisons in Fig. 2, Extended Data Fig. 1, 2, Extended Data Table 2, 3, and Supp. Table 3, 4. For tests that were performed both with and without sample size correction, the *p* value corresponding to the test with sample size correction was reported in the main text.

### Comparing neural activity across patients

To assess the progression of our feature over time, we compared the *R*^2^ time series progression across all patients. The 7-day moving average of the Δ*R*^2^ values for each patient was plotted with the average logistic regression decision threshold of each held out patient-fold. Then, for each clinical responder, we calculated the first day after DBS stimulation at which the moving Δ*R*^2^ average crossed the decision threshold derived from the fold in which that patient was left out (Extended Data Table 7). We then compared the decision threshold crossing day to the day clinical response was achieved, both normalized to the day of DBS activation (Fig. 4d).

### Comparing stimulation settings

We tested whether the change in neural predictability observed in responders was due to a fundamental difference in stimulation amplitude or pulse width. We plotted stimulation amplitude and pulse width for each hemisphere in responders and non-responders 14 days after stimulation began, and we tested significance between responders and non-responders in each hemisphere with Welch’s t-test (Supp. Table 7). Parameters not recoverable from device downloads were obtained from programming notes for three patients.

### Comparing neural activity and symptom severity

To assess the relationship between neural activity and symptom severity, we compared Y-BOCS scores with the average *R*^2^ values of the 14 days up to and including the day of assessment. We fit a mixed linear model (statsmodels 0.14.5) to correlate the clinical and *R*^2^ scores across patients to compensate for variable number of Y-BOCS timepoints captured per patient (N = 16 patients, n = 67 Y-BOCS scores). The model used the average 14-day *R*^2^ values as the dependent variable, Y-BOCS score as a fixed-effect predictor, and participants as the random intercepts. The model was fit using the restricted maximum likelihood (REML). The marginal *R*^2^ represents the variance of the fixed effects, and the conditional *R*^2^ represents the variance of both the fixed and random effects. Both values are reported in Fig. 2e.

To test how the Δ*R*^2^ feature at day 14 correlated with eventual symptom reduction at most recent follow-up, we plotted overall Y-BOCS reduction against day 14 Δ*R*^2^ for each patient and used a simple linear regression function (SciPy 1.16.3) to compute the correlation (Fig. 4i).

### Construction of circularized plots of daily 9 Hz power

To better visualize the 9 Hz power fluctuations over the course of the 24-hour period, we generated circularized polar plots of 9 Hz power on each day. We rounded each timestamp in our neural dataset up to the next multiple of 10 minutes so that our dataset had 144 unique times of day. Then, we took the average z-scored LFP power value at each of those 144 times of day for various subsets of the data (e.g., eventual responders during the pre-DBS state). We plotted this average value against time of day in polar plots, rotated so that midnight is aligned to 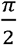 radians (upward on the polar plot), and time increases clockwise around the plot with a period of 24 hours. We also shaded the envelope representing one standard deviation in each direction of the values at each time.

### Oscillatory power of neural data

To analyze the periodicity of the pre-processed LFP power data, we computed the oscillatory power of the neural time series. For each day with a complete 4-day trailing window, we resampled the LFP time series into 30-minute averages, linearly interpolated and z-scored the window, and estimated the power spectral density using Welch’s method. We then compared the average power values across patients at the 6-, 12-, and 24-hour intervals between the pre-DBS vs. stable response (N=5) and pre-DBS vs. non-response (N=6) states. We tested significant differences from pre-DBS to stable response or non-response by using a paired t-test (Fig. 5c, Supp. Table 8).

### Acrophase calculation

We calculated the cosinor acrophase for each day of data in the dataset. We applied a sliding three-day window to each patient’s pre-processed neural time series, centered on the day of interest. We then fit a cosinor function of the following form to the data within each window (5):

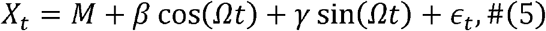

where *X*_*t*_ is the signal, *M* is the midline estimating statistic of rhythm (MESOR), *β* and *α* are respectively the cosine and sine coefficients, Ω is the frequency set to 24 hours (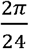 radians), and *ϵ*_*t*_ is the residual error. The parameters *M, β*, and *α* were estimated using a least squares approximation. We then calculated the following parameters of the fitted cosinor wave (6-8):

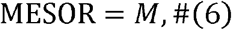

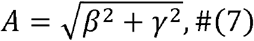

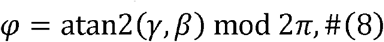

where atan2 is the four-quadrant inverse tangent function, and mod is the modulo function. The MESOR, amplitude (*A*), and acrophase (*φ*) in radians were assigned to the center day of each three-day sliding window. The acrophase can trivially be converted from radians to hours by multiplying by 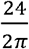.

The circular mean calculates the average of a collection of periodic values.^69^ Each angle is represented by a unit vector with orientation corresponding to that angle. We begin by converting these unit vectors to Cartesian coordinates (9, 10):

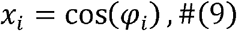

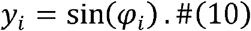

In this case, *φ*_*i*_ is the acrophase of day *i* = (1,…, *n*) in radians. Each unit vector is assigned a unit mass at the vector’s tip. The coordinates of the center of gravity *C* are then calculated (11, 12):

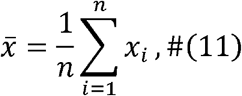

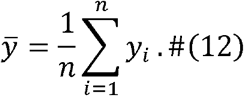

Finally, we find the angle of the center of gravity to get the circular mean angle (13):

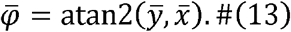

### Acrophase statistical methods

Because acrophase is a circular variable, we compared responder and non-responder distributions using the two-sample Mardia–Watson–Wheeler (uniform scores) test.^69^ The pooled angles from both samples are ranked around the unit circle from an arbitrary reference direction, each rank is mapped to a uniformly spaced angle, and the test statistic is based on the resultant vector length of the sum of all points belonging to each class. A large resultant indicates that class clusters in a preferred direction relative to the other. The statistic is approximately χ^2^-distributed for sufficiently large sample size, *N* = *m*+*n* > 14.^70^ In our case, we had *m* = 8 responders and *n* = 6 non-responders, so we calculated *p*-values via permutation testing (10,000 overall response-label shuffles) rather than relying on the asymptotic distribution.

### Acrophase comparisons

We plotted average acrophase for the pre-DBS period in each patient, separately for patients who eventually became stable clinical responders and non-responders. Circular plots aligned midnight to 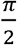 radians and displayed time of day progressing clockwise to represent acrophase. Average acrophase was calculated by taking the circular average of daily left-hemisphere acrophases in the pre-DBS period for each patient. The Mardia–Watson–Wheeler test was used to assess significance between the two groups (Supp. Table 9).

### Neural power differences by time of day

In order to quantify pre-DBS differences between responders and non-responders, we computed the average pre-DBS z-scored LFP power across all responders and non-responders (N=14), respectively, during each 10-minute interval throughout the day. We then plotted these averages with a one standard deviation envelope in either direction during the pre-DBS state. For each patient, we also averaged the pre-DBS z-scored LFP power for each 6-hour quadrant of the day (0:00-6:00, 6:00-12:00, 12:00-18:00, 18:00-24:00) and used a mixed linear model (statsmodels 0.14.5) to compare the average power 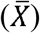 between responder groups across each time bin (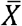 ~ group p time bin). The model was fit using the maximum likelihood (ML) with Powell’s optimization algorithm.^71^ Patients without data represented in each of the four quadrants were dropped. After applying FDR-Benjamini–Hochberg correction for multiple comparisons, we noted the time periods that had significant differences between eventual responders and non-responders (Fig. 5e, f, Supp. Table 10).

We then used a leave-one-patient-out framework to determine significant time periods within each patient-fold and fit a basic linear discriminant analysis model (scikit-learn 1.7.2). We used the average z-scored LFP power for each patient during the time of day represented by the significant time bins to classify the eventual response status of the left-out patient. If no time bins were deemed significant within a fold, all time bins were input as features. We compared the true classification performance to 10,000 shuffled eventual response-label runs and noted the value (Fig. 5g).

### Model comparison

We compared clinical state classification performance across three autoregressive model implementations: the *k*-lag linear autoregressive model (LinAR-*k* (zoned)^20^) we used previously, a new LinAR-*k* model trained with the novel 3-day sliding window paradigm described above instead of grouping the data by clinical state prior to training, and the LinAR-1 model we present in this work.

The LinAR-*k* models used significant lag terms up to 144 samples before the current value (*X*_1_ to *X*_144_), representing a full day. Significant terms were determined using a 5-fold cross-validation with an ordinary least squares regression model. For each fold of the cross-validation, significant lag terms (*p* < 0.05) were recorded. If the lag term was significant for more than half the folds, then the lag term was deemed significant. The models were run to predict LFP power values and calculate daily *R*^2^ values. The *R*^2^ values were used to calculate the Δ*R*^2^ feature, which was input as the feature for the same logistic regression framework described previously to classify symptom status. Prediction probabilities from each model were compared in a pairwise fashion using DeLong’s test. Results from the model performances and comparisons are shown in Supp. Table 5.

### Percept analysis application

To aid with consistent data analysis across DBS groups, we developed an application to help process, analyze, and visualize Medtronic Percept BrainSense Timeline data. The application and detailed documentation are available at https://github.com/ProvenzaLab/Percept_Data_Analysis_App. A reconstructed version of the final visualization panel is shown in Supp. Fig. 4.

The application was developed in Python using the PyQt family of packages. The analysis pipeline extracts neural power stored as BrainSense Timeline recordings from JSON files downloaded from the Percept device. Local field potential (LFP) data is then artifact-corrected, normalized, and run through an autoregressive model (AR) to predict LFP power values and calculate daily *R*^2^ values.

The GUI is easy to install as a local executable and can process multiple patients’ data at once. The user enters each patient’s initial DBS programming date, response status, and response date if applicable. A directory selection screen allows the user to easily select the parent directory containing the Medtronic Percept JSON files to be analyzed. One month of data is processed in ~30 seconds on a modern consumer-grade laptop. The GUI displays time-series plots for the raw LFP activity, AR-predicted neural activity, and daily *R*^2^ values for all patients input by the user. All computed variables and plots are downloadable in common data formats (i.e., CSV, PNG). The application does not require an internet connection to run and saves all data locally.

## Data availability

Deidentified participant level neural and clinical datasets that support the findings of this study are available upon request via the Data Archive for the BRAIN Initiative (DABI) registry https://dabi.loni.usc.edu/projects/AZB3H0HMN433/.

## Code availability

Custom code used to produce the results of this study is available at https://github.com/ProvenzaLab/PerceptPredictability.

## Acknowledgements

We are grateful to the patients and their families for their involvement in the research program. This research was supported by the National Institutes of Health (NIH) NINDS BRAIN Initiative via contract UH3 NS136631, NIH NIMH via contract R01 MH139889, the Brain and Behavior Research Foundation Young Investigator Award, and the McNair Foundation.

## Author contributions

N.R.P., S.A.S., and W.K.G. conceived of the study. N.R.P., S.A.S., R.R.H., and J.Z. conceptualized data analysis procedures, performed data analysis, interpreted data, and prepared figures and results, with support from T.M., R.B., A.K.A., V.A., T.M.F., S.R.H., W.K.G., E.A.S., A.B.P., and J.A.H. J.Z., S.S., A.K.A., S.R., V.R.G., and M.L. performed data collection in the clinic. N.R.P., A.B.P., R.R.H., J.Z., and S.A.S. contributed to conceptualization of the analytical study. A.B.P., N.R.P., and A.K.A. contributed to development of the autoregressive neural-predictability methodology and its statistical validation framework. J.Z., R.R.H., S.A.S., and N.R.P. wrote the first draft of the manuscript, and all authors contributed to the writing and revision of the manuscript. W.K.G., S.A.S., E.A.S., B.J.M., B.S., and B.M.K. performed the clinical care aspects of the study. S.A.S., D.L.P., K.A.K., and B.S. performed the study surgical procedures. N.R.P., S.A.S., W.K.G., and E.A.S. oversaw the collection of data, analysis, and manuscript completion.

## Competing interests

S.A.S. has been a consultant for Boston Scientific, Zimmer Biomet, Koh Young, Sensoria Therapeutics, Varian Medical Systems, Abbott and Neuropace and is co-founder of Motif Neurotech. W.K.G. receives royalties from Nview, LLC and OCDscales, LLC. E.A.S. receives direct funding from the International OCD Foundation as well as MHNTI for providing trainings on treating obsessive-compulsive disorder with psychotherapy. Furthermore, Dr. Storch co-founded Rethinking Behavioral Health which provides training and consultation in the treatment of obsessive-compulsive disorder and related conditions. He was a consultant for Brainsway and Biohaven Pharmaceuticals in the past 36 months. He owns stock options less than $5000 in NView (for distribution of the Y-BOCS and CY-BOCS) and receives royalties from OCD Scales LLC (for distribution of the Y-BOCS and CY-BOCS). He receives book royalties from Elsevier, Wiley, Oxford, American Psychological Association, Guildford, Springer, Routledge, and Jessica Kingsley.

## Ethics statement

Each patient gave fully informed consent to participate in this research study. The protocol was approved by the local institutional review boards at Baylor College of Medicine (IRB H-48392; H-56119) and the University of Utah (IRB 00169174).

## Notes

### Clinical Protocols

https://github.com/ProvenzaLab/PerceptPredictability

### Author Declarations

IRB H-48392; H-56119 of Baylor College of Medicine gave ethical approval for this work. IRB 00169174 of University of Utah gave ethical approval for this work.

