## Extended Figures for "Chronic intracranial neural dynamics forecast treatment response and therapeutic engagement during deep brain stimulation for OCD"

**Extended Data Figures**

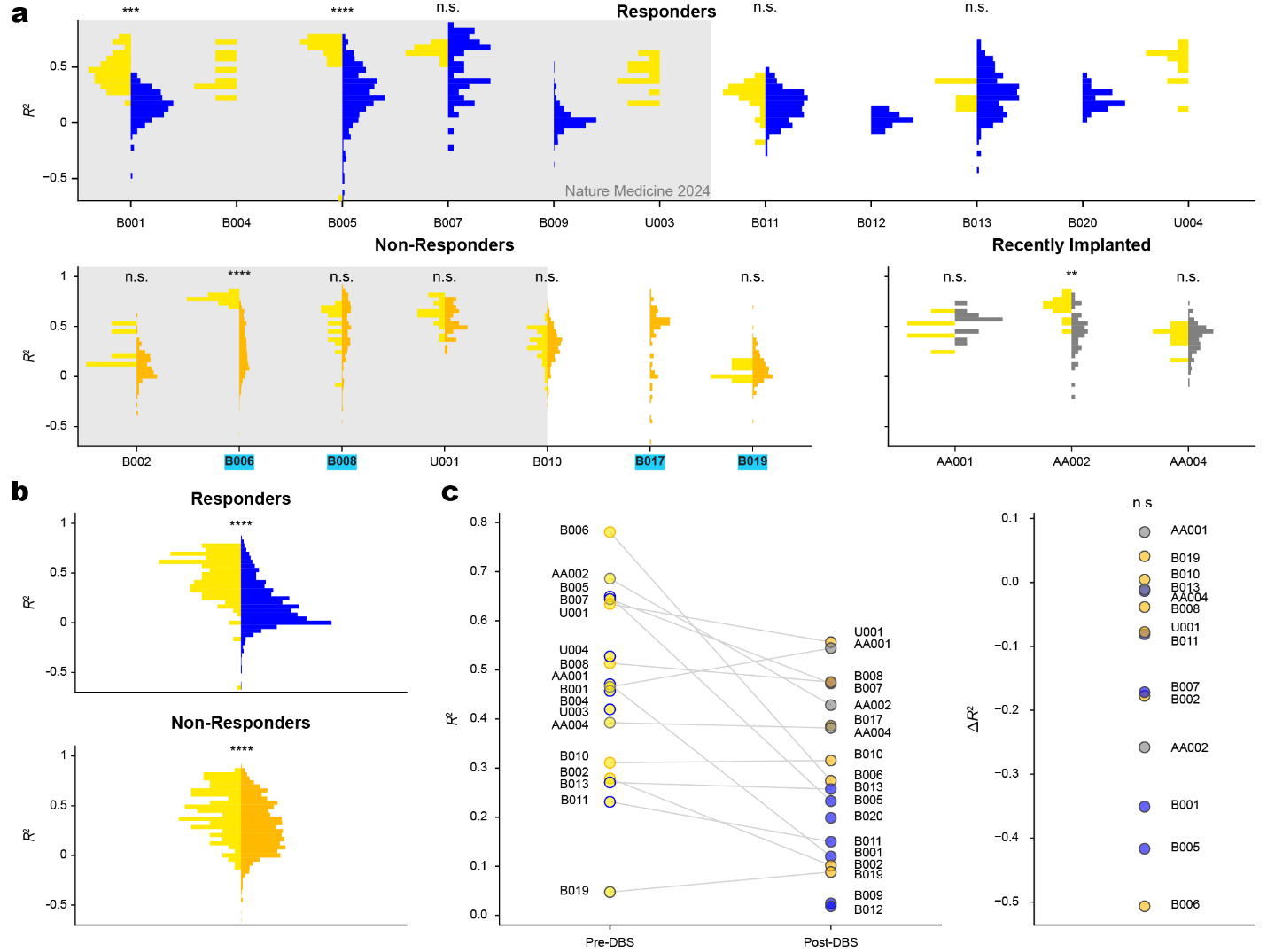

Extended Data Figure 1: **Right hemisphere neural predictability distributions. a,** Distributions of right hemisphere daily neural predictability during the pre-DBS and stable clinical states are displayed for individual patients (N=24). Patients included in the previous cohort (Provenza et al., 2024) are shown on a gray background. Pre-DBS values are shown in yellow, stable response values in blue, non-response values in orange, and values from recently implanted patients with undetermined outcomes in gray. Partial responder patient IDs are highlighted in light blue. Statistical comparisons for individual patients are reported in Supplementary Table 3. **b,** Pooled distributions of daily neural predictability $R^{2}$ for responders (N=13) and non-responders (N=7) during the pre-DBS and stable clinical states. Statistical results are reported in Supplementary Table 4. **c,** The clothesline plot displays the average $R^{2}$ during the pre-DBS and stable clinical states for all patients ($p=0.600$).

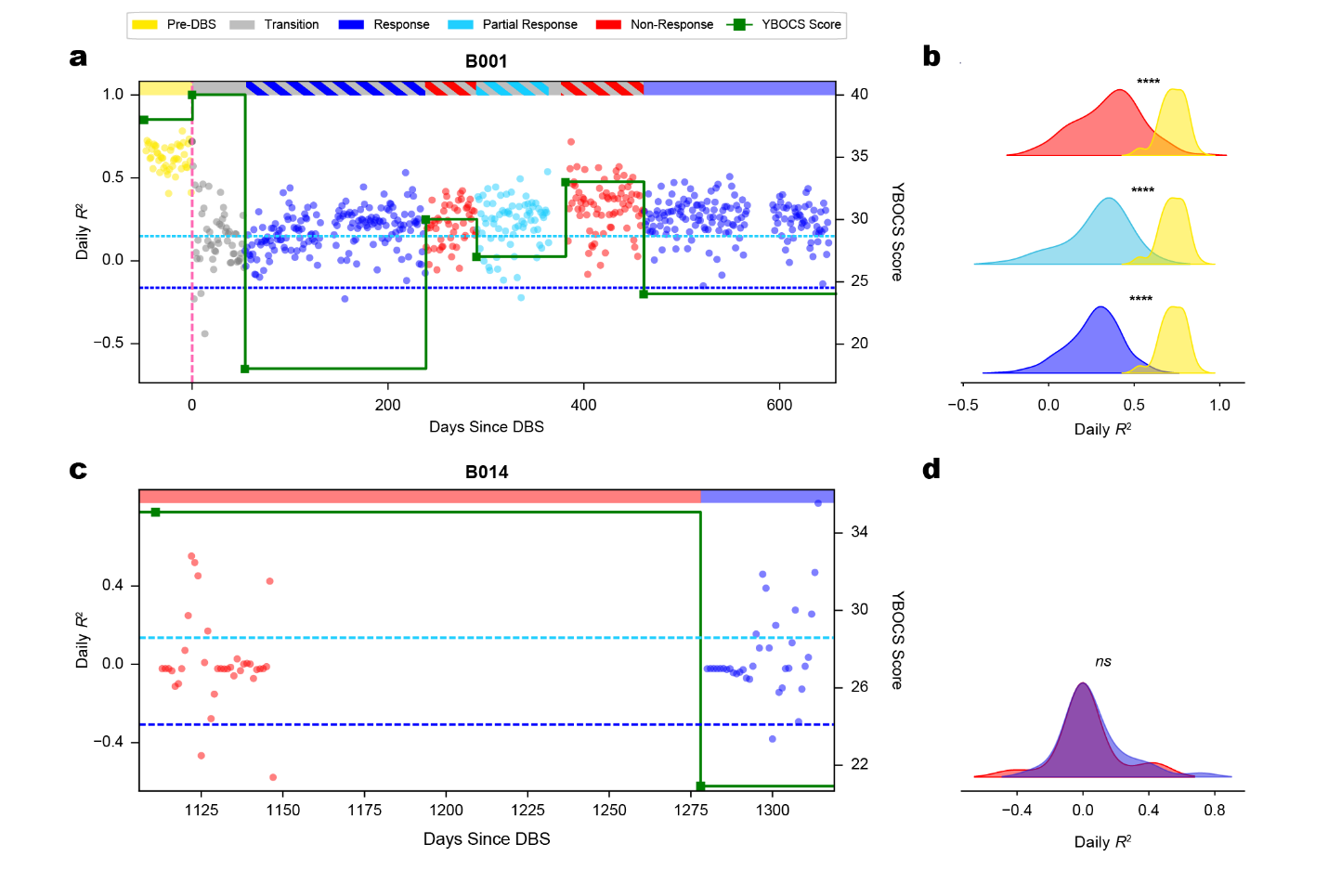

Extended Data Figure 2: **Neural predictability remains reduced during transient symptom fluctuations.** **a,** A responder patient (B001) experienced a period of temporary worsening of OCD symptoms (hashed red) during the transition period (hashed blue). Their $R^{2}$ time series is shown colored by the momentary clinical state based on the most recent Y-BOCS score. A colored bar at the top of the plot shows the true clinical state as defined in the Methods, with hash marks denoting periods with observed changes to symptoms during the transition state. The patient’s Y-BOCS scores are denoted with green boxes, with partial response and response Y-BOCS thresholds for this patient shown by dashed lines in light blue and blue, respectively. **b,** Distributions and comparisons between pre-DBS and acute non-response, partial response, and response periods are shown. Corresponding $p$ values are $p=3.11\times{10}^{-10}, 3.10\times{10}^{-11}, 1.29\times{10}^{-14}$ (top to bottom). **c,** A patient (B014) experienced a period of sustained symptom relapse (292 days) approximately 1.5 years after stable response was achieved. In total, only 74 days of neural data were recorded, but captured periods of relapse and subsequent stable response. Their $R^{2}$ time series and Y-BOCS scores are shown with the same color scheme as **a, b**. **d,** The $R^{2}$ distributions of the non-response and response states are shown ($p=0.675$). All statistical tests used Welch’s two-sample t-test using ESS-adjusted sample sizes with Benjamini–Hochberg FDR correction.

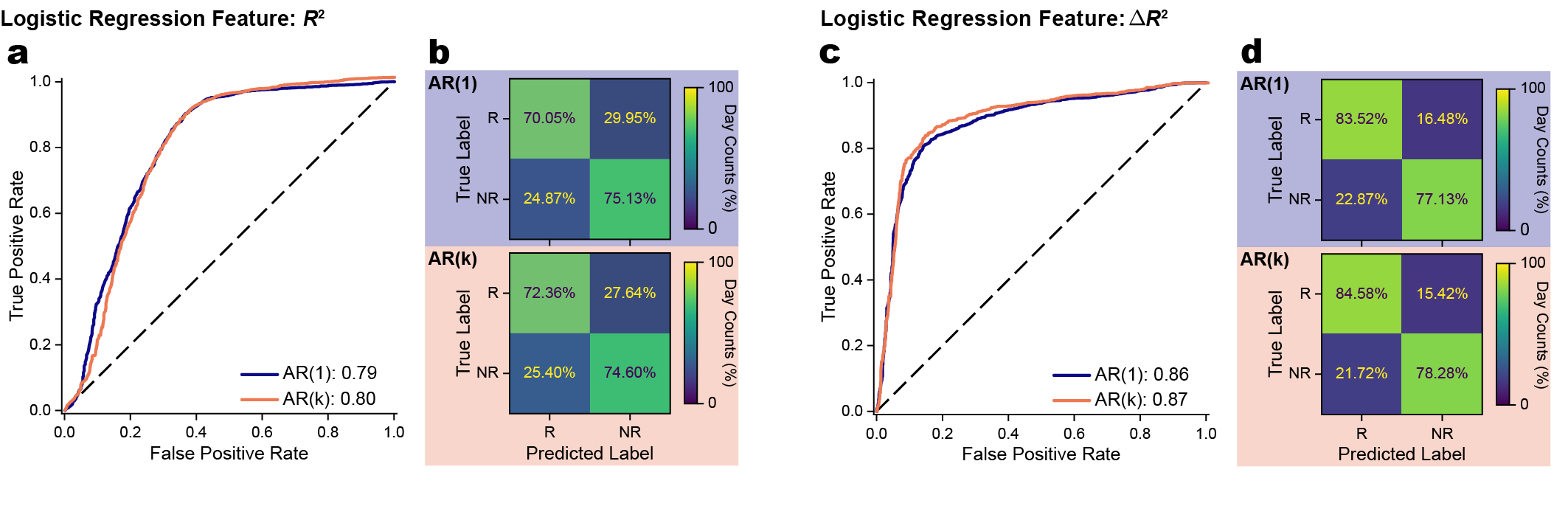

Extended Data Figure 3: **An autoregressive model with** $\boldsymbol{k}$**-lags performs similarly to a simpler model.** Daily $R^{2}$ values were calculated with a LinAR-1 and LinAR-$k$ model. $R^{2}$ values were used to predict daily symptom status using a logistic regression with a leave-one-patient-out cross-validation. **a,** ROC curves display the model performance for the LinAR-1 and LinAR-$k$ model. **b,** Confusion matrices for the LinAR-1 and LinAR-$k$ models. The confusion matrices display the percentage of days predicted within each state. **c, d,** Analogous plots are shown with $\Delta R^{2}$ as the input feature. Model performance statistics are reported in Extended Data Table 5.

**Extended Data Tables**

| **Patient ID** | **Gender** | **Age at time of surgery (years)** | **Follow-up**  **duration (months)** | **Responder status** | **Baseline Y-BOCS**  **(Y-BOCS II) score** | **Final Y-BOCS**  **(Y-BOCS II) score** | **# of Y-BOCS Assessments** | **Left\|Right recording duration (days) (^20^)** |
| --- | --- | --- | --- | --- | --- | --- | --- | --- |
| AA001 | F | 16-20 | 10 | Undet. | 37 (43) | 32 (34) | 15 | 141\|27 (-) |
| AA002 | M | 26-30 | 12 | Undet. | 37 (41) | 35 (38) | 15 | 136\|65 (-) |
| AA004 | M | 56-60 | 6 | Undet. | 36 (38) | 26 (29) | 6 | 95\|94 (-) |
| B001 | F | 41-45 | 24 | R | 38 (39) | 24 (26) | 7 | 650\|650 (150\|150) |
| B002 | F | 31-35 | 2 | NR | 40 (44) | 39 (42) | 2 | 99\|210 (99\|210) |
| B004 | M | 31-35 | 38 | R | 34 (34) | 11 (13) | 6 | 544\|11 (411\|11) |
| B005 | M | 16-20 | 34 | R | 34 (42) | 10 (11) | 5 | 482\|482 (380\|380) |
| B006 | F | 51-55 | 43 | PR | 39 (43) | 37 (38) | 9 | 829\|917 (344\|432) |
| B007 | M | 21-25 | 25 | R | 33 (35) | 28 (30) | 5 | 15\|97 (15\|59) |
| B008 | F | 31-35 | 37 | PR | 35 (36) | 26 (27) | 4 | 382\|361 (138\|117) |
| B009 | F | 31-35 | 53 | R | 37 (47) | 0 (0) | 16 | 303\|303 (120\|120) |
| B010 | F | 21-25 | 12 | NR | 30 (32) | 31 (33) | 9 | 329\|329 (24\|24) |
| B011 | M | 46-50 | 13 | R | 34 (37) | 10 (11) | 6 | 220\|220 (-) |
| B012 | F | 36-40 | 59 | R | 39 (49) | 16 (18) | 15 | 37\|37 (-) |
| B013 | M | 21-25 | 11 | R | 40 (43) | 18 (19) | 7 | 231\|231 (-) |
| B014 | M | 36-40 | 45 | R | 36 (45) | 26 (29) | 17 | 74\|0 (-) |
| B015 | F | 26-30 | 32 | R | 40 (45) | 36 (37) | 17 | 74\|0 (-) |
| B017 | M | 31-35 | 38 | PR | 38 (46) | 26 (28) | 15 | 89\|87 (-) |
| B019 | M | 36-40 | 18 | PR | 40 (42) | 30 (32) | 9 | 164\|164 (-) |
| B020 | M | 36-40 | 65 | R | 38 (42) | 25 (26) | 17 | 37\|37 (-) |
| U001 | F | 31-35 | 2 | NR | 32 (-) | 32 (-) | 3 | 69\|69 (69\|69) |
| U002 | M | 21-25 | 4 | R | 30 (-) | 17 (-) | 3 | 15\|29 (16\|30) |
| U003 | F | 21-25 | 10 | R | 34 (-) | 21 (-) | 3 | 96\|96 (29\|29) |
| U004 | F | 41-45 | 1 | R | 28 (-) | 20 (-) | 6 | 15\|15 (-) |

Extended Data Table 1: **Patient demographics, clinical outcomes, and duration of continuous neural recordings.** Gender was based on self-report. Follow-up duration reflects the interval between initial surgery and the most recent clinical visit or Y-BOCS scale assessment. Responder status was determined as stable responder, non-responder, or undetermined based on criteria described in the Methods. Patients B012, B014-B017, and B020 were implanted with the Percept device after undergoing a battery replacement, so the initial Y-BOCS and Y-BOCS II scores are from the timepoint of the initial surgery. The duration of continuous recording days included in our previous study^20^ is also listed. R, Responder; NR, Non-Responder; PR, Partial Responder; Undet., Undetermined; Y-BOCS, Yale–Brown obsessive-compulsive scale.

| **Patient ID** | **N_eff 1_** | **N_eff 2_** | **DF** | **T-stat** | $\boldsymbol{p}$**-value** | **95% CI** | **Hedges G** | **Hedges G CI** |
| --- | --- | --- | --- | --- | --- | --- | --- | --- |
| AA001 | 5.64 | 13.9 | 17.1 | 3.77 | 1.52e-3 | (0.117, 0.415) | 1.34 | (0.313, 2.36) |
| AA002 | 4.91 | 29.7 | 7.70 | 3.47 | 8.95e-3 | (0.0761, 0.384) | 1.19 | (0.213, 2.16) |
| AA004 | 3.49 | 24.2 | 7.93 | -2.50 | 0.0374 | (-0.235, -9.13e-3) | -0.757 | (-1.86, 0.351) |
| B001 | 12.5 | 55.2 | 22.8 | 13.7 | 8.49e-13 | (0.317, 0.430) | 3.39 | (2.55, 4.22) |
| B002 | 3.17 | 23.3 | 2.46 | -0.0571 | 0.959 | (-0.472, 0.457) | -0.0446 | (-1.18, 1.09) |
| B004 | 4.39 | 54.5 | 6.95 | 9.85 | 2.49e-05 | (0.381, 0.622) | 2.44 | (1.39, 3.50) |
| B005 | 8.16 | 75.5 | 9.46 | 4.35 | 1.64e-3 | (0.123, 0.384) | 1.39 | (0.645, 2.14) |
| B006 | 7.16 | 56.6 | 25.1 | 2.81 | 9.45e-3 | (0.0150, 0.0972) | 0.527 | (-0.246, 1.30) |
| B007 | - | - | - | - | - | - | - | - |
| B008 | 3.88 | 27.9 | 4.89 | 0.146 | 0.889 | (-0.266, 0.298) | 0.0571 | (-0.978, 1.09) |
| B009 | - | - | - | - | - | - | - | - |
| B010 | 12.9 | 106 | 14.5 | 0.638 | 0.534 | (-0.0797, 0.147) | 0.199 | (-0.375, 0.773) |
| B011 | 11.4 | 48.8 | 11.2 | 4.77 | 5.56e-04 | (0.208, 0.562) | 2.56 | (1.77, 3.34) |
| B012 | - | - | - | - | - | - | - | - |
| B013 | 4.84 | 55.6 | 6.09 | 3.47 | 0.0129 | (0.0642, 0.366) | 1.03 | (0.0979, 1.97) |
| B014 | - | - | - | - | - | - | - | - |
| B015 | - | - | - | - | - | - | - | - |
| B017 | - | - | - | - | - | - | - | - |
| B019 | 4.09 | 23.0 | 8.82 | 0.591 | 0.569 | (-0.104, 0.178) | 0.192 | (-0.830, 1.21) |
| B020 | - | - | - | - | - | - | - | - |
| U001 | 7.70 | 10.4 | 15.8 | 2.40 | 0.0292 | (0.0149, 0.244) | 1.02 | (0.0699, 1.97) |
| U002 | - | - | - | - | - | - | - | - |
| U003 | - | - | - | - | - | - | - | - |
| U004 | - | - | - | - | - | - | - | - |

Extended Data Table 2: **Statistics for comparison of left hemisphere pre-DBS versus stable state periods in each patient with sample size correction.** The statistical test used was the Welch’s t-test. Clinical responders are highlighted in light blue, non-responders in light orange, and patients whose clinical state has not yet been determined in gray. Patients without statistics lacked either pre- or post-DBS data. DF, degrees of freedom; CI, confidence interval.

|  |  | **Status** | **N_1_** | **N_2_** | **DF** | **T-stat** | $\boldsymbol{p}$**-value** | **95% CI** | **Hedges G** | **Hedges G CI** |
| --- | --- | --- | --- | --- | --- | --- | --- | --- | --- | --- |
| **Uncorrected Sample Size** | $R^{2}$ | Responder | 183 | 1685 | 219 | 20.5 | 7.21e-53 | (0.279, 0.338) | 1.66 | (1.50, 1.82) |
|  |  | Non-Responder | 115 | 1699 | 132 | -1.44 | 0.151 | (-0.0786, 0.0123) | -0.134 | (-0.322, 0.0553) |
|  |  | Undetermined | 30 | 327 | 36.3 | 4.81 | 2.63e-05 | (0.103, 0.252) | 0.820 | (0.442, 1.20) |
|  | $\Delta R^{2}$ | Responder | 183 | 1220 | 279 | 25.6 | 3.28e-75 | (0.321, 0.374) | 1.70 | (1.53, 1.87) |
|  |  | Non-Responder | 115 | 1663 | 137 | 2.74 | 6.95e-3 | (0.0114, 0.0702) | 0.230 | (0.0413, 0.419) |
|  |  | Undetermined | 30 | 327 | 70.2 | 6.62 | 6.06e-9 | (0.108, 0.202) | 0.625 | (0.249, 1.00) |
| **Corrected Sample Size** | $R^{2}$ | Responder | 61.3 | 357 | 80.1 | 11.6 | 9.20e-19 | (0.255, 0.361) | 1.66 | (1.36, 1.95) |
|  |  | Non-Responder | 38.9 | 257 | 51.4 | -0.806 | 0.424 | (-0.115, 0.0494) | -0.134 | (-0.470, 0.203) |
|  |  | Undetermined | 14.0 | 67.8 | 20.7 | 3.09 | 5.60e-3 | (0.0579, 0.297) | 0.822 | (0.239, 1.41) |
|  | $\Delta R^{2}$ | Responder | 61.3 | 289 | 106 | 14.3 | 2.07e-26 | (0.299, 0.396) | 1.71 | (1.41, 2.02) |
|  |  | Non-Responder | 38.9 | 247 | 55.7 | 1.51 | 0.136 | (-0.0132, 0.0948) | 0.232 | (-0.105, 0.570) |
|  |  | Undetermined | 14.0 | 67.8 | 52.0 | 3.75 | 4.50e-4 | (0.0720, 0.238) | 0.645 | (0.0671, 1.22) |

Extended Data Table 3: **Statistics for comparison of left hemisphere pre-DBS versus stable state periods’** $\boldsymbol{R}^{\boldsymbol{2}}$ **pooled across all patients.** The statistical test used was the Welch’s t-test. Abbreviations are the same as in Extended Data Table 2.

|  |  | **Window (days)** | **AUROC** | **BA** | **TPR** | **TNR** | **EER Threshold**  **(mean / min / max)** |
| --- | --- | --- | --- | --- | --- | --- | --- |
| **Weighted by Day** | $\boldsymbol{R}^{\boldsymbol{2}}$ | 1 | 0.785 | 0.697 | 0.636 | 0.758 | - / - / - |
|  |  | 3 | 0.814 | 0.705 | 0.636 | 0.774 | - \| - / - |
|  |  | 5 | 0.825 | 0.705 | 0.632 | 0.777 | - \| - / - |
|  |  | 7 | 0.829 | 0.700 | 0.620 | 0.781 | - \| - / - |
|  |  | 14 | 0.834 | 0.696 | 0.616 | 0.776 | - \| - / - |
|  |  | 21 | 0.837 | 0.688 | 0.599 | 0.776 | - \| - / - |
|  |  | 28 | 0.840 | 0.680 | 0.580 | 0.779 | - \| - / - |
|  | $\boldsymbol{\Delta}\boldsymbol{R}^{\boldsymbol{2}}$ | 1 | 0.857 | 0.796 | 0.757 | 0.835 | - \| - / - |
|  |  | 3 | 0.913 | 0.853 | 0.812 | 0.893 | - \| - / - |
|  |  | 5 | 0.928 | 0.875 | 0.833 | 0.916 | - \| - / - |
|  |  | 7 | 0.937 | 0.884 | 0.847 | 0.921 | - \| - / - |
|  |  | 14 | 0.949 | 0.899 | 0.872 | 0.927 | - \| - / - |
|  |  | 21 | 0.956 | 0.908 | 0.892 | 0.925 | - \| - / - |
|  |  | 28 | 0.962 | 0.913 | 0.922 | 0.903 | - \| - / - |
| **Weighted by Patient** | $\boldsymbol{R}^{\boldsymbol{2}}$ | 1 | 0.738 | 0.693 | 0.716 | 0.722 | 0.576 / 0.553 / 0.600 |
|  |  | 3 | 0.762 | 0.707 | 0.732 | 0.740 | 0.614 / 0.591 / 0.637 |
|  |  | 5 | 0.776 | 0.725 | 0.747 | 0.756 | 0.625 / 0.601 / 0.651 |
|  |  | 7 | 0.782 | 0.731 | 0.744 | 0.767 | 0.631 / 0.607 / 0.659 |
|  |  | 14 | 0.788 | 0.732 | 0.747 | 0.767 | 0.643 / 0.618 / 0.669 |
|  |  | 21 | 0.791 | 0.726 | 0.735 | 0.765 | 0.654 / 0.627 / 0.679 |
|  |  | 28 | 0.793 | 0.720 | 0.720 | 0.766 | 0.655 / 0.627 / 0.685 |
|  | $\boldsymbol{\Delta}\boldsymbol{R}^{\boldsymbol{2}}$ | 1 | 0.858 | 0.789 | 0.782 | 0.797 | 0.492 / 0.457 / 0.566 |
|  |  | 3 | 0.916 | 0.845 | 0.833 | 0.857 | 0.478 / 0.431 / 0.568 |
|  |  | 5 | 0.932 | 0.859 | 0.849 | 0.869 | 0.472 / 0.384 / 0.587 |
|  |  | 7 | 0.940 | 0.868 | 0.861 | 0.875 | 0.471 / 0.365 / 0.588 |
|  |  | 14 | 0.950 | 0.886 | 0.880 | 0.891 | 0.431 / 0.353 / 0.568 |
|  |  | 21 | 0.959 | 0.906 | 0.898 | 0.914 | 0.411 / 0.338 / 0.499 |
|  |  | 28 | 0.966 | 0.924 | 0.927 | 0.921 | 0.361 / 0.313 / 0.453 |

Extended Data Table 4: **LinAR-1 symptom-burdened classification regression stats across averaged windows.** Regression performance statistics are shown for averaged $R^{2}$ and $\Delta R^{2}$ values across increasing window widths. Statistics were computed aggregated across the entire dataset (weighted by day) and by taking the mean of per-patient averages such that each patient had equal weight (weighted by patient). EER thresholds were only calculated at the patient level for each held-out patient and set at the equal error rate of the training patients out-of-fold predictions. AUROC, area under the receiver operating characteristic curve; BA, balanced accuracy; TPR, true positive rate; TNR, true negative rate, EER, equal error rate.

| **Hem.** | **Feature** | **Label** | **AUROC** | **BA** | **TPR** | **TNR** |
| --- | --- | --- | --- | --- | --- | --- |
| **Left** | $R^{2}$ | True | 0.784 | 0.693 | 0.621 | 0.766 |
|  |  | Shuffled | 0.496 | 0.499 | 0.499 | 0.499 |
|  |  | $p$ (random) | 1.00E-04 | 1.00E-04 | 0.082 | 2.00E-04 |
|  |  | $p$ (circular) | 1.00E-04 | 1.00E-04 | 1.00E-04 | 0.997 |
|  | $\Delta R^{2}$ | True | 0.857 | 0.796 | 0.757 | 0.835 |
|  |  | Shuffled | 0.496 | 0.500 | 0.500 | 0.500 |
|  |  | $p$ (random) | 1.00E-04 | 1.00E-04 | 4.00E-04 | 1.00E-04 |
|  | Averaged $\Delta R^{2}$ | True | 0.949 | 0.899 | 0.872 | 0.927 |
|  |  | Shuffled | 0.496 | 0.500 | 0.501 | 0.499 |
|  |  | $p$ (random) | 1.00E-04 | 1.00E-04 | 2.00E-04 | 1.00E-04 |
| **Right** | $R^{2}$ | True | 0.619 | 0.613 | 0.680 | 0.545 |
|  |  | Shuffled | 0.496 | 0.500 | 0.499 | 0.500 |
|  |  | $p$ (random) | 1.00E-04 | 1.00E-04 | 3.50E-03 | 0.434 |
|  |  | $p$ (circular) | 1.00E-04 | 1.00E-04 | 3.00E-04 | 0.730 |
|  | $\Delta R^{2}$ | True | 0.074 | 0.143 | 0.149 | 0.137 |
|  |  | Shuffled | 0.497 | 0.498 | 0.495 | 0.501 |
|  |  | $p$ (random) | 1.00 | 1.00 | 0.995 | 1.00 |
|  | Averaged $\Delta R^{2}$ | True | 0.014 | 0.076 | 0.056 | 0.095 |
|  |  | Shuffled | 0.496 | 0.497 | 0.497 | 0.496 |
|  |  | $p$ (random) | 1.00 | 1.00 | 1.00 | 1.00 |

Extended Data Table 5: **LinAR-1 symptom-burdened classification regression statistics.** Statistics are shown for both hemispheres using three feature sets: $R^{2}$, $\Delta R^{2}$, and 14-day averaged $\Delta R^{2}$ values. The near-zero right-hemisphere AUROC values for $\Delta R^{2}$ and averaged $\Delta R^{2}$ do not indicate an inverted discriminant. Instead, the responder and non-responder distributions overlap so closely that the sign of the fitted coefficient depends on which patient is held out, resulting in seemingly poor test performance for that patient. Circular p values were calculated only for the daily feature since the delta and averaged features were each limited to a single class (i.e., either responder or non-responder). Abbreviations are the same as in Extended Data Table 4.

| **Model** | **Feature** | **Data** | **AUROC** | **BA** | **TPR** | **TNR** |
| --- | --- | --- | --- | --- | --- | --- |
| **LinAR-1** | $R^{2}$ | Old | 0.795 | 0.663 | 0.553 | 0.774 |
|  |  | All | 0.784 | 0.693 | 0.621 | 0.766 |
|  | $\Delta R^{2}$ | Old | 0.708 | 0.665 | 0.512 | 0.818 |
|  |  | All | 0.857 | 0.796 | 0.757 | 0.835 |
|  | Averaged $\Delta R^{2}$ | Old | 0.579 | 0.589 | 0.400 | 0.777 |
|  |  | All | 0.949 | 0.899 | 0.872 | 0.927 |
| **LinAR-**$\boldsymbol{k}$ | $R^{2}$ | Old | 0.816 | 0.693 | 0.555 | 0.830 |
|  |  | All | 0.801 | 0.704 | 0.619 | 0.789 |
|  | $\Delta R^{2}$ | Old | 0.668 | 0.656 | 0.493 | 0.820 |
|  |  | All | 0.868 | 0.805 | 0.730 | 0.880 |
|  | Averaged $\Delta R^{2}$ | Old | 0.609 | 0.616 | 0.404 | 0.828 |
|  |  | All | 0.953 | 0.903 | 0.853 | 0.952 |
| **LinAR-**$\boldsymbol{k}$ **(zoned)** | $R^{2}$ | Old | 0.865 | 0.745 | 0.686 | 0.804 |
|  |  | All | 0.813 | 0.742 | 0.710 | 0.773 |
|  | $\Delta R^{2}$ | Old | 0.608 | 0.640 | 0.512 | 0.767 |
|  |  | All | 0.747 | 0.702 | 0.707 | 0.696 |
|  | Averaged $\Delta R^{2}$ | Old | 0.532 | 0.569 | 0.401 | 0.736 |
|  |  | All | 0.810 | 0.745 | 0.797 | 0.693 |

Extended Data Table 6: **Logistic regression model daily clinical state classification performance.** We evaluated performance of three models, LinAR-1, LinAR-$k$, and the previously established LinAR-$k$ (zoned)^20^ model across two datasets (the original^20^ dataset (old) and all available data) using three feature sets: $R^{2}$, $\Delta R^{2}$, and 14-day averaged $\Delta R^{2}$ values. Abbreviations are the same as in Extended Data Table 4.

| **Patient ID left out** | **Response status** | **Raw accuracy** | **Logistic regression decision threshold** |
| --- | --- | --- | --- |
| B001 | Responder | 0.982 | -0.155 |
| B002 | Non-Responder | 0.886 | -0.182 |
| B004 | Responder | 0.969 | -0.141 |
| B005 | Responder | 0.551 | -0.220 |
| B006 | Non-Responder | 0.903 | -0.187 |
| B008 | Non-Responder | 0.712 | -0.165 |
| B010 | Non-Responder | 0.834 | -0.179 |
| B011 | Responder | 0.981 | -0.154 |
| B013 | Responder | 0.428 | -0.220 |
| B019 | Non-Responder | 0.794 | -0.173 |
| U001 | Non-Responder | 0.651 | -0.164 |
| OVERALL | - | 0.802 | - |

Extended Data Table 7: **Per-fold logistic regression performance and threshold for model fit on the** $\boldsymbol{\Delta}\boldsymbol{R}^{\boldsymbol{2}}$ **feature without smoothing.** The decision threshold for each held-out patient was set at the equal error rate of the training patients' out-of-fold predictions.
