## Supplemental Figures for "Chronic intracranial neural dynamics forecast treatment response and therapeutic engagement during deep brain stimulation for OCD"

**Supplementary Figures**

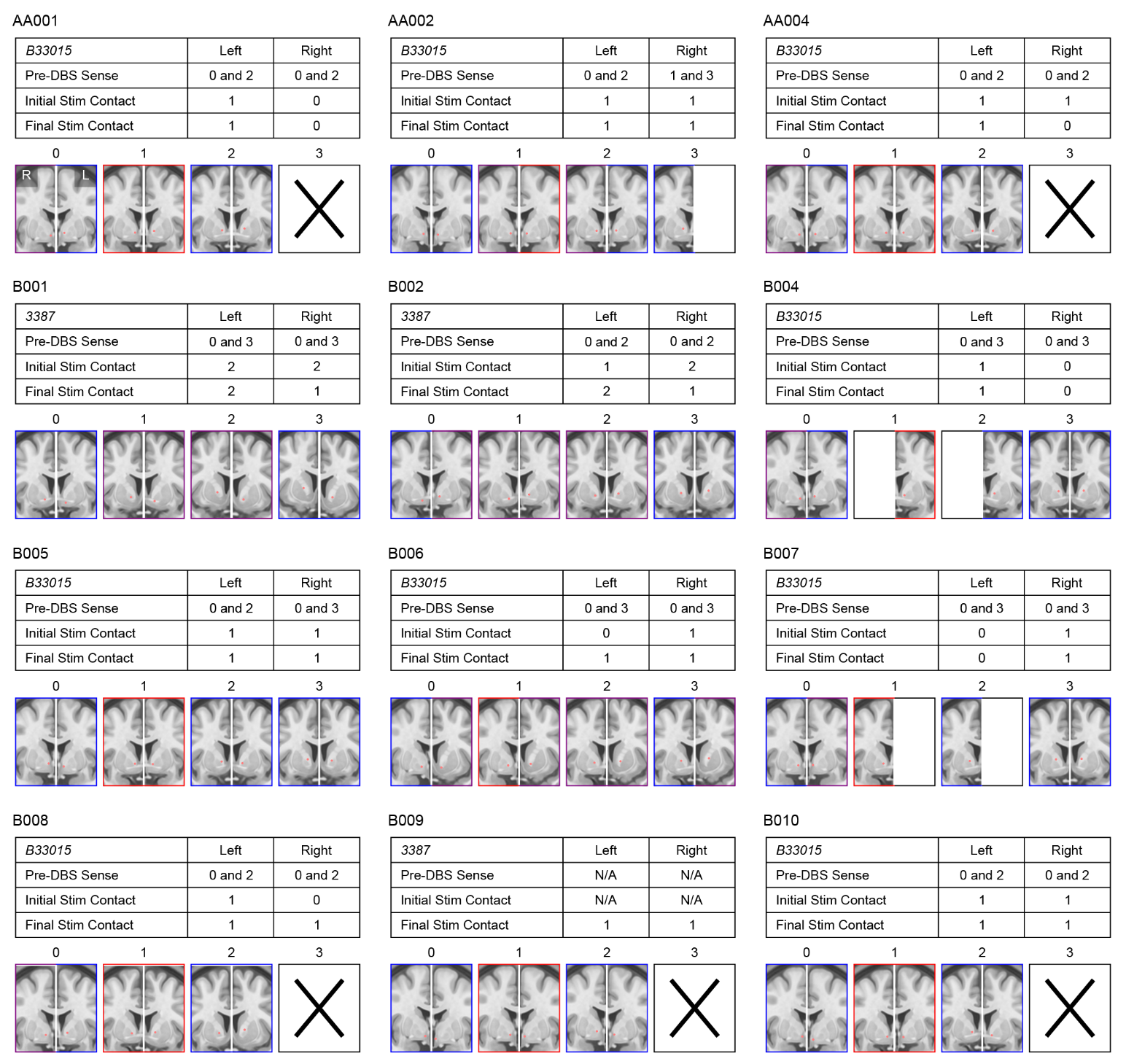

Supplementary Figure 1: **Stimulating and sensing contact locations and configurations throughout data collection.** Coronal slices show stimulating and sensing contact locations for patients AA001 through B010 with treatment-resistant OCD, bilaterally implanted in the VC/VS or BNST. Contacts are denoted by red dots and shown in MNI152 space. Slice borders indicate whether contacts were used for stimulation (red), sensing (blue), or both (purple) over the course of data collection; unused contacts are not shown. Pre-DBS sensing and initial and final (i.e., at the last data sample collected) post-DBS stimulation configurations are listed for each patient. For directional Medtronic SenSight leads, the most anterior segment was selected to represent each segmented contact level. Stimulation and sensing configurations were extracted from Percept session JSON downloads and programming notes. Displayed contact coordinates are recorded in Supp. Table 1.

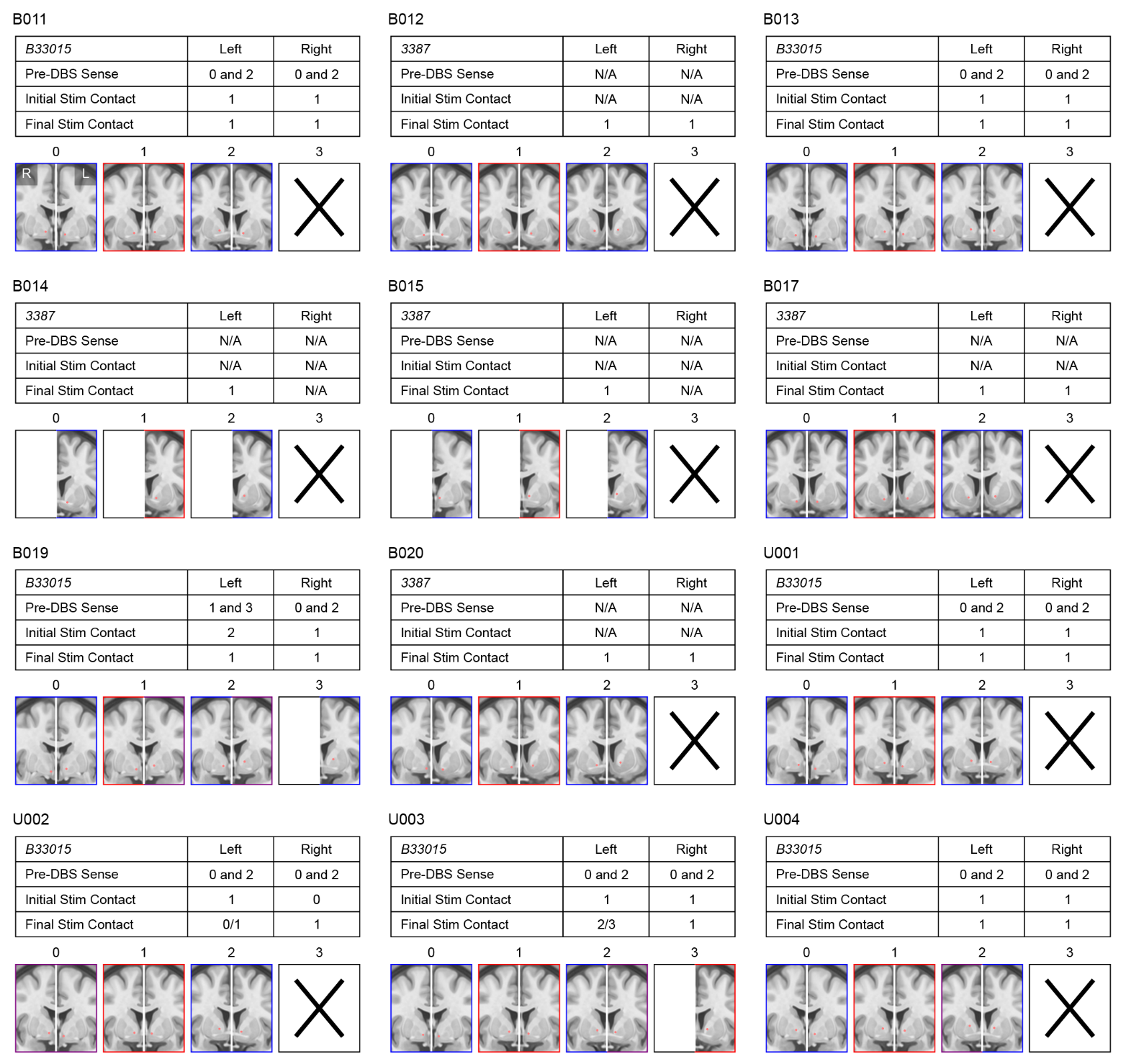

Supplementary Figure 2: **Analogous plots to Supplementary Figure 1 for patients B011 through U004.** Contact configurations containing a slash (e.g., 0/1) denote double monopolar stimulation.

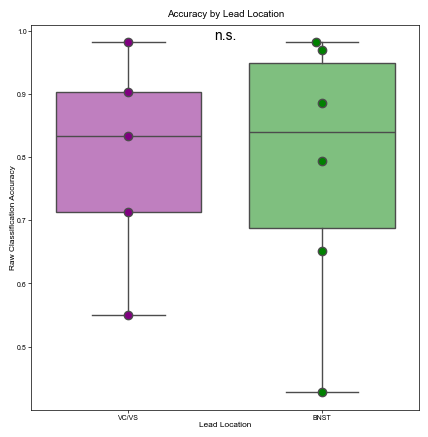

Supplementary Figure 3: **Classification accuracy did not differ by lead trajectory.** Box plots show per-patient raw classification accuracy from the logistic regression model trained on the $\Delta R^{2}$ feature for patients implanted with the VC/VS (N=5) and BNST trajectories (N=6). A Welch’s two-tailed t-test indicated no significant difference ($p=0.923$).

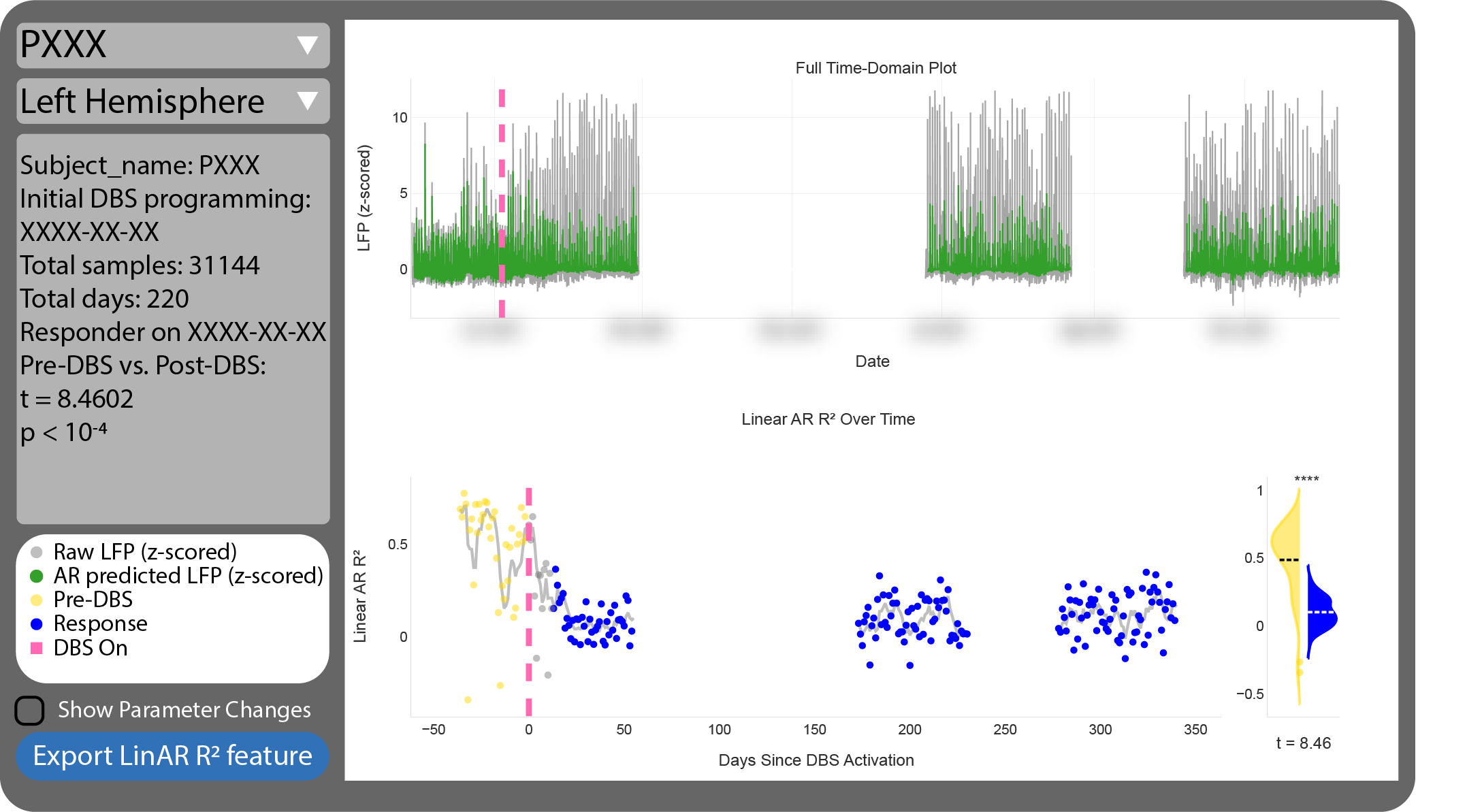

Supplementary Figure 4: **A user-friendly application to evaluate neural predictability.** A reconstructed screenshot of the streamlined application to analyze neural predictability. The interface displays a graph of the z-scored 9 Hz LFP time series (gray) and the autoregressive model's predicted LFP time series (green). The bottom plot shows a scatter plot of the $R^{2}$ time series colored by clinical status and a 2-week moving average of the $R^{2}$ values (gray). An associated violin plot shows the distributions of the pre-DBS (if available) and stable clinical $R^{2}$ values. The corresponding $t$ value and $p$ value (denoted by the stars) are shown as well. The interface displays the current patient ID, hemisphere, basic stats, and a legend on the left. An option to display vertical lines on the graphs corresponding to DBS parameter changes and export the $R^{2}$ features to a .csv file is also available.

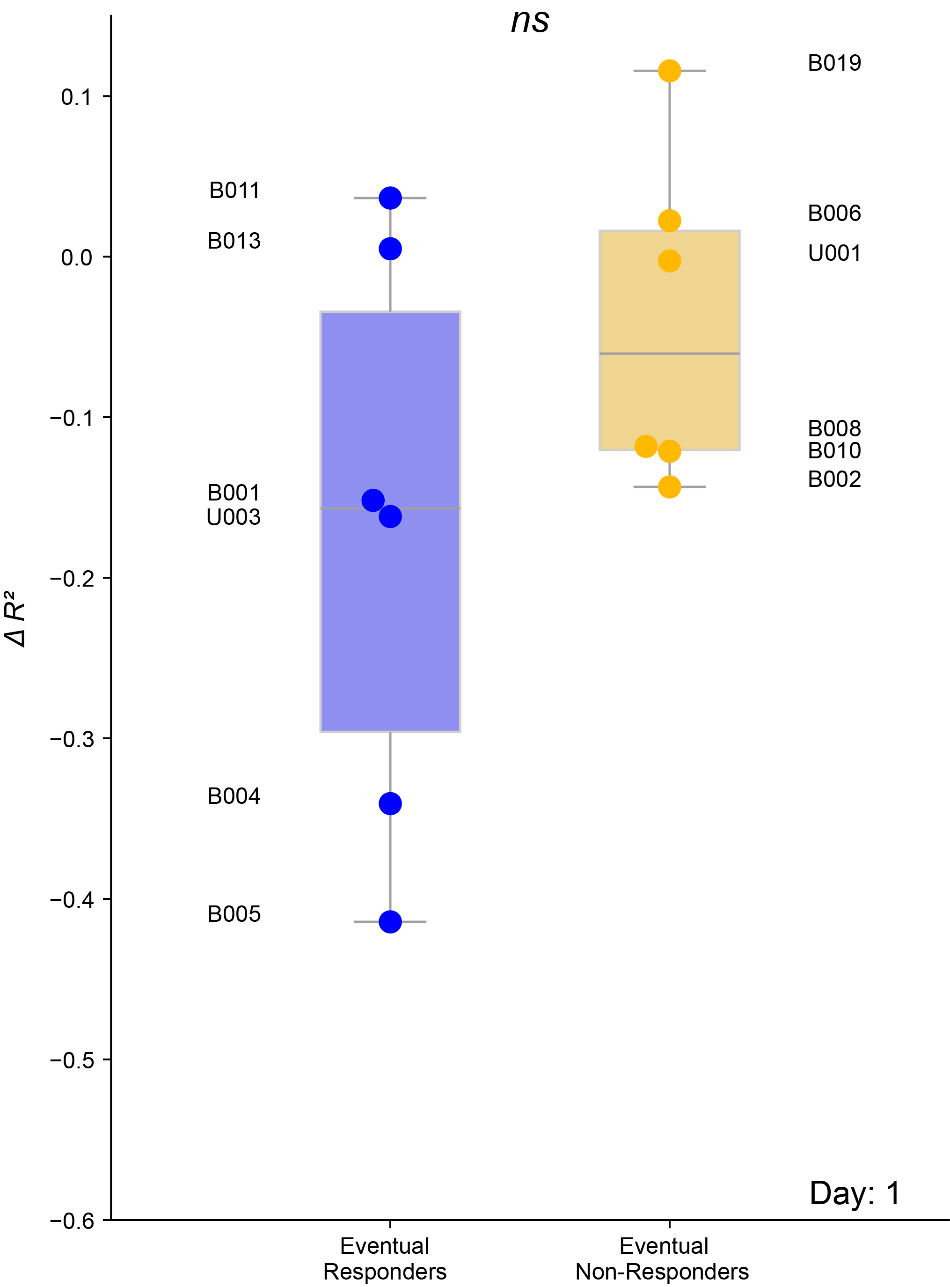

Supplementary Video 1: **Video of** $\boldsymbol{\Delta}\boldsymbol{R}^{\boldsymbol{2}}$ **values over time.** Distributions of eventual responders and non-responders' average $\Delta R^{2}$ values on each day are shown over time after DBS activation. Significance between the two distributions was calculated using Welch's t-test with Benjamini-Hochberg FDR correction across the first 500 days after DBS activation.

**Supplementary Tables**

|  |  | **Left** | | | | **Right** | | | |
| --- | --- | --- | --- | --- | --- | --- | --- | --- | --- |
| **Patient ID** | **Lead Model** | **0** | **1** | **2** | **3** | **0** | **1** | **2** | **3** |
| AA001 | B33015 | (-8.9, -0.7, -2.7) | (-11.1, 1.4, -0.7) | (-13.1, 2.9, 1.7) | Not Used | (5.6, -0.1, -5.3) | (8.2, 2, -2.9) | (10.6, 3.3, -0.6) | Not Used |
| AA002 | B33015 | (-6.2, -2.1, -5.3) | (-8.1, -0.5, -3.3) | (-9.8, 0.7, -0.8) | Not Used | (6.4, -1, -5.1) | (8.1, 0.8, -3.3) | (9.7, 2, -1) | (11.4, 2.6, 1.5) |
| AA004 | B33015 | (-9.1, -0.7, -4.6) | (-10.9, 1.4, -3) | (-12.7, 3, -0.9) | Not Used | (6.5, -0.8, -5.1) | (8.3, 1.7, -3.9) | (10.1, 3.7, -2.1) | Not Used |
| B001 | 3387 | (-10.6, 2.1, -9.4) | (-12, 3.9, -6.9) | (-13.3, 5.7, -4.4) | (-14.7, 7.6, -2.1) | (11.9, 4.8, -5.4) | (13.5, 6.5, -2.9) | (15, 8.2, -0.5) | (16.6, 10, 1.9) |
| B002 | 3387 | (-9.8, 0.4, -4.4) | (-11.8, 2.1, -2) | (-13.7, 3.7, 0.4) | (-15.5, 5.2, 2.7) | (8.8, 2.8, -8.5) | (10.6, 4.6, -5.8) | (12.4, 6.2, -3.2) | (14.3, 7.9, -0.7) |
| B004 | B33015 | (-8.6, 0.9, -3.9) | (-10.7, 2.6, -1.6) | (-12.8, 3.7, 1) | (-14.9, 4.2, 4) | (6.8, 0.2, -5) | (8.5, 2.1, -2.7) | (10.2, 3.4, 0.1) | (11.9, 4.1, 3.3) |
| B005 | B33015 | (-8.4, 0.3, -4.8) | (-10.1, 2.4, -3.6) | (-12, 3.9, -1.7) | (-13.9, 4.9, 0.6) | (7.4, 1.6, -2.8) | (9.1, 3.7, -1.6) | (10.7, 5.3, 0.1) | (12.3, 6.5, 2.4) |
| B006 | B33015 | (-11.5, 7.1, -7.6) | (-13.2, 8.3, -4.8) | (-14.9, 8.9, -1.8) | (-16.8, 9, 1.6) | (10, -0.4, -4.6) | (11.4, 1.5, -2.4) | (12.9, 2.8, 0.1) | (14.6, 3.5, 3.2) |
| B007 | B33015 | (-6.2, 0.7, -2.9) | (-8.2, 2.5, -0.8) | (-10.3, 3.9, 1.7) | (-12.5, 4.6, 4.6) | (9, 1.5, -2.8) | (11, 3, -0.4) | (13, 4, 2.3) | (15.1, 4.4, 5.4) |
| B008 | B33015 | (-11.6, 1.7, -0.3) | (-13.6, 4.2, 0.4) | (-15.4, 6.4, 1.8) | Not Used | (9.9, 0.7, -3.3) | (11.5, 3.3, -2.2) | (13.1, 5.6, -0.6) | Not Used |
| B009 | 3387 | (-3.1, -2.2, -7.7) | (-5.4, -1.2, -5.6) | (-7.7, -0.1, -3.5) | Not Used | (6.2, -0.2, -8.5) | (8.4, 0.4, -5.7) | (10.4, 1, -3) | Not Used |
| B010 | B33015 | (-5.7, -1.7, -8.2) | (-8, 0.5, -6.4) | (-10.2, 2.2, -4.4) | Not Used | (5.9, 0.4, -6.4) | (7.9, 2.4, -4.3) | (9.9, 4.1, -2) | (11.8, 5.3, 0.7) |
| B011 | B33015 | (-7.5, -0.8, -5.3) | (-9.4, 1.1, -3.6) | (-11.4, 2.4, -1.5) | Not Used | (9.3, 0.6, -5.2) | (11, 2.5, -3.5) | (12.8, 3.9, -1.4) | Not Used |
| B012 | 3387 | (-11.1, 2.9, -5.6) | (-12.5, 4.4, -3.2) | (-13.9, 5.8, -0.8) | Not Used | (8.9, 3.7, -5.9) | (10.8, 5.5, -3.9) | (12.6, 7.2, -1.9) | Not Used |
| B013 | B33015 | (-7.3, -2.8, -5.9) | (-9, -0.6, -4.1) | (-10.6, 1, -1.9) | Not Used | (9.8, -1.3, -5.1) | (11.4, 0.4, -2.8) | (13.1, 1.4, -0.1) | Not Used |
| B014 | 3387 | (-11.3, 3.6, -5.8) | (-13.1, 4.4, -3.1) | (-15, 5.1, -0.4) | Not Used | Not Used | Not Used | Not Used | Not Used |
| B015 | 3387 | (-7.4, 0.2, -3) | (-9.3, 1.9, -0.7) | (-11.2, 3.6, 1.7) | Not Used | Not Used | Not Used | Not Used | Not Used |
| B017 | 3387 | (-10.2, 5.1, -9) | (-12, 6.4, -6.8) | (-13.8, 7.7, -4.5) | Not Used | (10.3, 5.7, -8.6) | (11.9, 7.2, -6.4) | (13.4, 8.7, -4.2) | Not Used |
| B019 | B33015 | (-10.2, -2.6, -3.7) | (-11.9, -0.4, -1.7) | (-13.7, 1.1, 0.6) | (-15.7, 1.8, 3.1) | (5.9, -2.9, -7.7) | (7.8, -0.6, -5.5) | (9.5, 0.9, -3) | Not Used |
| B020 | 3387 | (-11.2, 5.3, -8.6) | (-12.6, 6.4, -5.7) | (-14, 7.6, -3) | Not Used | (6.3, 1.1, -8.2) | (8.1, 2.3, -5.7) | (9.9, 3.5, -3.3) | Not Used |
| U001 | B33015 | (-6.7, -1, -6.5) | (-8.5, 1.2, -4.7) | (-10.2, 2.8, -2.3) | Not Used | (9.1, 0.3, -5.2) | (10.8, 1.9, -2.8) | (12.7, 2.7, -0.1) | Not Used |
| U002 | B33015 | (-6.8, -0.8, -5.9) | (-8.6, 1.4, -4.2) | (-10.4, 2.9, -1.9) | Not Used | (9.1, 0.5, -5.3) | (10.8, 2.1, -2.8) | (12.6, 2.9, -0.1) | Not Used |
| U003 | B33015 | (-6.8, -0.8, -6.2) | (-8.5, 1.4, -4.3) | (-10.2, 2.9, -1.9) | (-12, 3.9, 1.1) | (8.9, 0.6, -5.3) | (10.5, 2.2, -2.7) | (12.3, 3.1, 0.1) | Not Used |
| U004 | B33015 | (-9.4, -1.3, -3.6) | (-11, 0.9, -0.7) | (-12.6, 2.3, 2.6) | Not Used | (11.4, -0.2, -1.9) | (13.1, 1.9, 0.9) | (14.9, 3.3, 4) | Not Used |

Supplementary Table 1: **MNI coordinates of stimulating and sensing contacts.** Each patient received bilateral DBS leads targeting the VC/VS or VC/BNST. Lead models included Medtronic SenSight B33015 and Medtronic 3387 (legacy) leads. Contact 0 is the most ventral, and contact 3 is the most dorsal. For directional Medtronic SenSight B33015 leads, the most anterior segment was selected to represent each segmented contact level. MNI, Montreal Neurological Institute.

| **Patient ID** | **N_1_** | **N_2_** | **DF** | **T-stat** | $\boldsymbol{p}$**-value** | **95% CI** | **Hedges G** | **Hedges G CI** |
| --- | --- | --- | --- | --- | --- | --- | --- | --- |
| AA001 | 7 | 128 | 9.49 | 6.39 | 9.99e-5 | (0.173, 0.360) | 1.26 | (0.489, 2.03) |
| AA002 | 15 | 114 | 24.6 | 6.26 | 1.61e-6 | (0.154, 0.306) | 1.20 | (0.649, 1.76) |
| AA004 | 8 | 85 | 16.6 | -4.13 | 7.43e-4 | (-0.184, -0.0594) | -0.766 | (-1.49, -0.0392) |
| B001 | 47 | 164 | 106 | 25.5 | 3.60e-47 | (0.344, 0.402) | 3.46 | (2.99, 3.92) |
| B002 | 5 | 70 | 4.27 | -0.0727 | 0.945 | (-0.280, 0.266) | -0.0463 | (-0.944, 0.852) |
| B004 | 8 | 322 | 9.02 | 15.0 | 1.12e-7 | (0.426, 0.577) | 2.43 | (1.71, 3.15) |
| B005 | 43 | 385 | 55.9 | 9.97 | 5.20e-14 | (0.203, 0.304) | 1.40 | (1.08, 1.73) |
| B006 | 13 | 787 | 15.9 | 5.19 | 9.02e-5 | (0.0332, 0.0790) | 0.512 | (-0.0359, 1.06) |
| B007 | - | - | - | - | - | - | - | - |
| B008 | 18 | 351 | 21.1 | 0.342 | 0.736 | (-0.0810, 0.113) | 0.0577 | (-0.415, 0.530) |
| B009 | - | - | - | - | - | - | - | - |
| B010 | 50 | 271 | 65.2 | 1.22 | 0.225 | (-0.0214, 0.0891) | 0.199 | (-0.102, 0.500) |
| B011 | 36 | 162 | 37.5 | 8.46 | 3.20e-10 | (0.293, 0.477) | 2.58 | (2.14, 3.02) |
| B012 | - | - | - | - | - | - | - | - |
| B013 | 5 | 187 | 4.67 | 3.82 | 0.0141 | (0.0671, 0.364) | 1.03 | (0.136, 1.92) |
| B014 | - | - | - | - | - | - | - | - |
| B015 | - | - | - | - | - | - | - | - |
| B017 | - | - | - | - | - | - | - | - |
| B019 | 6 | 141 | 6.97 | 0.874 | 0.411 | (-0.0627, 0.136) | 0.190 | (-0.624, 1.00) |
| B020 | - | - | - | - | - | - | - | - |
| U001 | 23 | 43 | 61.7 | 4.58 | 2.33e-5 | (0.0729, 0.186) | 1.03 | (0.497, 1.56) |
| U002 | - | - | - | - | - | - | - | - |
| U003 | - | - | - | - | - | - | - | - |
| U004 | - | - | - | - | - | - | - | - |

Supplementary Table 2: **Statistics for comparison of left hemisphere pre-DBS versus stable state periods in each patient without sample size correction.** The statistical test used was the Welch’s t-test. Clinical responders are highlighted in light blue, non-responders in light orange, and patients whose clinical state has not yet been determined in gray. Patients without statistics lacked either pre- or post-DBS data. DF, degrees of freedom; CI, confidence interval.

| **Patient ID** | **N_eff 1_** | **N_eff 2_** | **DF** | **T-stat** | $\boldsymbol{p}$**-value** | **95% CI** | **Hedges G** | **Hedges G CI** |
| --- | --- | --- | --- | --- | --- | --- | --- | --- |
| AA001 | 5.19 | 5.29 | 8.27 | -0.642 | 0.538 | (-0.264, 0.149) | -0.360 | (-1.47, 0.752) |
| AA002 | 5.19 | 10.4 | 13.4 | 3.12 | 7.90e-3 | (0.0794, 0.434) | 1.29 | (0.201, 2.39) |
| AA004 | 4.10 | 16.6 | 5.98 | -0.703 | 0.508 | (-0.206, 0.114) | -0.317 | (-1.36, 0.724) |
| B001 | 8.53 | 65.4 | 9.16 | 5.79 | 2.46e-04 | (0.190, 0.433) | 2.28 | (1.48, 3.08) |
| B002 | 2.25 | 50.4 | 1.31 | 1.35 | 0.364 | (-0.799, 1.16) | 1.25 | (-0.0877, 2.59) |
| B004 | - | - | - | - | - | - | - | - |
| B005 | 23.9 | 54.7 | 48.3 | 7.03 | 6.39e-09 | (0.280, 0.504) | 1.64 | (1.10, 2.18) |
| B006 | 6.52 | 39.4 | 43.9 | 13.3 | 5.31e-17 | (0.429, 0.582) | 2.37 | (1.42, 3.32) |
| B007 | 6.24 | 9.93 | 10.2 | 1.91 | 0.0839 | (-0.0260, 0.351) | 0.741 | (-0.240, 1.72) |
| B008 | 11.0 | 121 | 12.9 | 0.565 | 0.582 | (-0.108, 0.184) | 0.150 | (-0.464, 0.763) |
| B009 | - | - | - | - | - | - | - | - |
| B010 | 13.5 | 62.6 | 20.0 | -0.140 | 0.890 | (-0.106, 0.0930) | -0.0386 | (-0.620, 0.543) |
| B011 | 11.1 | 42.6 | 15.8 | 1.69 | 0.111 | (-0.209, 0.184) | 0.562 | (-0.0969, 1.22) |
| B012 | - | - | - | - | - | - | - | - |
| B013 | 3.99 | 42.8 | 4.840 | -0.0516 | 0.961 | (-0.176, 0.169) | -0.0170 | (-1.03, 0.992) |
| B014 | - | - | - | - | - | - | - | - |
| B015 | - | - | - | - | - | - | - | - |
| B017 | - | - | - | - | - | - | - | - |
| B019 | 2.37 | 21.6 | 2.69 | -1.27 | 0.302 | (-0.330, 0.150) | -0.502 | (-1.80, 0.801) |
| B020 | - | - | - | - | - | - | - | - |
| U001 | 6.52 | 15.3 | 12.8 | 1.27 | 0.228 | (-0.0520, 0.199) | 0.525 | (-0.371, 1.42) |
| U002 | - | - | - | - | - | - | - | - |
| U003 | - | - | - | - | - | - | - | - |
| U004 | - | - | - | - | - | - | - | - |

Supplementary Table 3: **Statistics for comparison of right hemisphere pre-DBS versus stable state** $\boldsymbol{R}^{\boldsymbol{2}}$**values in each patient with sample size correction.** The statistical test used was the Welch’s t-test. Clinical responders are highlighted in light blue, non-responders in light orange, and patients whose clinical state has not yet been determined in gray. Patients without statistics lacked either pre- or post-DBS data. DF, degrees of freedom; CI, confidence interval.

|  |  | **Status** | **N_1_** | **N_2_** | **DF** | **T-stat** | $\boldsymbol{p}$**-value** | **95% CI** | **Hedges G** | **Hedges G CI** |
| --- | --- | --- | --- | --- | --- | --- | --- | --- | --- | --- |
| **Uncorrected Sample Size** | $\boldsymbol{R}^{\boldsymbol{2}}$ | Responder | 183 | 1320 | 233 | 16.5 | 7.30e-41 | (0.251, 0.320) | 1.322 | (1.16, 1.48) |
|  |  | Non-Responder | 115 | 1870 | 128 | 6.17 | 8.33e-9 | (0.0982, 0.191) | 0.599 | (0.410, 0.789) |
|  |  | Undetermined | 30 | 123 | 43.0 | 2.95 | 0.00519 | (0.0338, 0.181) | 0.612 | (0.209, 1.01) |
|  | $\boldsymbol{\Delta}\boldsymbol{R}^{\boldsymbol{2}}$ | Responder | 183 | 960 | 397 | 16.1 | 3.49e-45 | (0.206, 0.263) | 0.939 | (0.776, 1.10) |
|  |  | Non-Responder | 115 | 1837 | 193 | 16.6 | 3.81e-39 | (0.229, 0.290) | 0.824 | (0.634, 1.01) |
|  |  | Undetermined | 30 | 123 | 92.1 | 2.17 | 0.0324 | (0.00538, 0.120) | 0.300 | (-0.0986, 0.698) |
| **Corrected Sample Size** | $\boldsymbol{R}^{\boldsymbol{2}}$ | Responder | 69.0 | 325 | 97.5 | 9.81 | 3.34e-16 | (0.228, 0.343) | 1.32 | (1.04, 1.59) |
|  |  | Non-Responder | 42.2 | 318 | 52.4 | 3.62 | 6.60e-4 | (0.0645, 0.225) | 0.598 | (0.275, 0.921) |
|  |  | Undetermined | 14.5 | 32.3 | 25.1 | 1.91 | 0.0683 | (-0.00869, 0.223) | 0.602 | (-0.0197, 1.22) |
|  | $\boldsymbol{\Delta}\boldsymbol{R}^{\boldsymbol{2}}$ | Responder | 69.0 | 215 | 191 | 8.90 | 4.16e-16 | (0.183, 0.287) | 0.965 | (0.683, 1.25) |
|  |  | Non-Responder | 42.2 | 310 | 107 | 8.94 | 1.25e-14 | (0.202, 0.317) | 0.844 | (0.517, 1.17) |
|  |  | Undetermined | 14.5 | 32.3 | 44.1 | 1.27 | 0.211 | (-0.0370, 0.163) | 0.312 | (-0.301, 0.924) |

Supplementary Table 4: **Statistics for comparison of right hemisphere pre-DBS versus stable state periods’** $\boldsymbol{R}^{\boldsymbol{2}}$ **pooled across all patients.** The statistical test used was the Welch’s t-test. Abbreviations are the same as in Supplementary Table 3.

| **Feature** | **Other model** | $\boldsymbol{p}$ | $\boldsymbol{z}$ |
| --- | --- | --- | --- |
| $\boldsymbol{R}^{\boldsymbol{2}}$ | LinAR-$k$ | 0.476 | -0.712 |
|  | LinAR-$k$ (zoned) | 0.00592 | 2.752 |
| $\boldsymbol{\Delta}\boldsymbol{R}^{\boldsymbol{2}}$ | LinAR-$k$ | 3.00e-06 | 4.671 |
|  | LinAR-$k$ (zoned) | 1.76e-29 | 11.274 |
| **Averaged** $\boldsymbol{\Delta}\boldsymbol{R}^{\boldsymbol{2}}$ | LinAR-$k$ | 0.0585 | 1.892 |
|  | LinAR-$k$ (zoned) | 5.60e-65 | 17.022 |

Supplementary Table 5: **LinAR-1 model performance comparison with Delong’s Test.** Regression performance comparison between the LinAR-1 model and LinAR-$k$ models for all three $R^{2}$ features using Delong's test to compare prediction probabilities.

| **Patient ID** | **Response status** | **Average** $\boldsymbol{\Delta R}^{\boldsymbol{2}}$ **(days 1-14)** |
| --- | --- | --- |
| B001 | Responder | -0.486 |
| B002 | Non-Responder | 0.0420 |
| B004 | Responder | -0.379 |
| B005 | Responder | -0.472 |
| B006 | Non-Responder | 0.0601 |
| B008 | Non-Responder | -0.198 |
| B010 | Non-Responder | 0.0102 |
| B011 | Responder | -0.225 |
| B013 | Responder | -0.166 |
| B019 | Non-Responder | -0.0305 |
| U001 | Non-Responder | -0.0473 |
| U003 | Responder | -0.0127 |

Supplementary Table 6: **Per-patient average** $\boldsymbol{\Delta R}^{\boldsymbol{2}}$ **values of days 1-14 after DBS activation.**

| **Patient ID** | **Response status** | **Left amp. (mA)** | **Right amp. (mA)** | **Left pulse width (µs)** | **Right pulse width (µs)** |
| --- | --- | --- | --- | --- | --- |
| B001 | Responder | 3.0 | 4.0 | 120 | 90 |
| B002 | Non-Responder | 3.5 | 3.8 | 120 | 120 |
| B004 | Responder | 4.0 | 4.5 | 120 | 90 |
| B005 | Responder | 3.0 | 3.0 | 90 | 90 |
| B006 | Non-Responder | 3.5 | 3.5 | 90 | 120 |
| B008 | Non-Responder | 3.5 | 4.0 | 90 | 120 |
| B010 | Non-Responder | 3.5 | 3.5 | 90 | 90 |
| B011 | Responder | 2.0 | 2.0 | 90 | 90 |
| B013 | Responder | 4.0 | 4.0 | 90 | 90 |
| B019 | Non-Responder | 4.0 | 3.5 | 90 | 90 |
| U001 | Non-Responder | 3.5 | 3.5 | 90 | 90 |

Supplementary Table 7: **Patient stimulation parameters on day 14 after DBS activation.** Each patient’s stimulation amplitude (mA) and pulse width (µs) are shown for both hemispheres. In patients with adjustable amplitude configurations, the upper limit was selected. Values were verified via programming notes. Amp., amplitude.

|  |  | **Pre-DBS** | | | **Stable State** | | |
| --- | --- | --- | --- | --- | --- | --- | --- |
| **Patient ID** | **Response Status** | **PSD 6-**  **hour comp.** | **PSD 12-**  **hour comp.** | **PSD 24-**  **hour comp.** | **PSD 6-**  **hour comp.** | **PSD 12-**  **hour comp.** | **PSD 24-**  **hour comp.** |
| B001 | R | 0.0285 | 0.0426 | 0.292 | 0.0248 | 0.0409 | 0.162 |
| B002 | NR | 0.0106 | 0.0696 | 0.265 | 0.0145 | 0.0260 | 0.159 |
| B004 | R | 0.0169 | 0.0809 | 0.311 | 0.0164 | 0.0303 | 0.0857 |
| B005 | R | 0.0220 | 0.0767 | 0.117 | 0.0146 | 0.0359 | 0.0857 |
| B006 | NR | 0.00903 | 0.0336 | 0.267 | 0.0224 | 0.0522 | 0.125 |
| B008 | NR | 0.0143 | 0.0339 | 0.0835 | 0.0151 | 0.0325 | 0.0837 |
| B010 | NR | 0.0144 | 0.0429 | 0.106 | 0.0163 | 0.0342 | 0.0931 |
| B011 | R | 0.0115 | 0.0563 | 0.183 | 0.0157 | 0.0223 | 0.0600 |
| B013 | R | 0.0113 | 0.0691 | 0.161 | 0.0153 | 0.0368 | 0.0942 |
| B019 | NR | 0.0188 | 0.0270 | 0.0218 | 0.0148 | 0.0318 | 0.0647 |
| U001 | NR | 0.0270 | 0.0484 | 0.156 | 0.0281 | 0.0270 | 0.0956 |

Supplementary Table 8: **PSD power of different components during pre-DBS and stable post-DBS clinical states for all patients.** Average Welch PSD power (au.) of the 6-, 12-, and 24-hour component during the pre-DBS and stable clinical states for each patient. PSD, power spectral density; comp., component.

| **Patient ID** | **Response Status** | **Average Left Acrophase (rad.)** | **Average Right Acrophase (rad.)** |
| --- | --- | --- | --- |
| AA001 | Undefined | 0.762 | 0.178 |
| AA002 | Undefined | 4.30 | 4.41 |
| AA004 | Undefined | 0.833 | 0.804 |
| B001 | Responder | 3.81 | 4.71 |
| B002 | Non-Responder | 3.48 | 3.27 |
| B004 | Responder | 0.596 | 0.453 |
| B005 | Responder | 0.624 | 0.468 |
| B006 | Non-Responder | 1.06 | 1.10 |
| B007 | Responder | 0.0680 | 0.515 |
| B008 | Non-Responder | 1.48 | 1.16 |
| B010 | Non-Responder | 4.35 | 4.69 |
| B011 | Responder | 0.617 | 5.65 |
| B013 | Responder | 0.165 | 5.73 |
| B019 | Non-Responder | 2.69 | 4.50 |
| U001 | Non-Responder | 3.85 | 3.90 |
| U003 | Responder | 5.63 | 4.86 |
| U004 | Responder | 0.726 | 0.970 |

Supplementary Table 9: **Average pre-DBS acrophase (in radians) per hemisphere across patients.** Values span from 0 radians at the start of the day to 2$\pi$ at its end and are defined to increase linearly over the 24-hour cycle.

| **Patient ID** | **Response**  **Status** | **0:00-6:00**  **Power (a.u.)** | **6:00-12:00**  **Power (a.u.)** | **12:00-18:00**  **Power (a.u.)** | **18:00-24:00**  **Power (a.u.)** |
| --- | --- | --- | --- | --- | --- |
| B001 | Responder | -0.632 | 0.0399 | 0.670 | -0.0858 |
| B002 | Non-Responder | -0.478 | 0.246 | 0.377 | -0.210 |
| B004 | Responder | 0.972 | -0.337 | -0.527 | -0.0964 |
| B005 | Responder | 0.586 | -0.202 | -0.252 | -0.132 |
| B006 | Non-Responder | 0.907 | 0.106 | -0.509 | -0.465 |
| B007 | Responder | 0.455 | -0.411 | 0.0947 | -0.149 |
| B008 | Non-Responder | 0.0902 | 0.0743 | -0.0797 | -0.0776 |
| B010 | Non-Responder | -0.129 | 0.0138 | 0.0845 | 0.0312 |
| B011 | Responder | 0.806 | -0.219 | -0.419 | -0.162 |
| B013 | Responder | 0.409 | -0.412 | -0.304 | 0.336 |
| B019 | Non-Responder | 0.00181 | 0.123 | 0.133 | -0.230 |
| U001 | Non-Responder | -0.367 | -0.175 | 0.584 | -0.0592 |
| U003 | Responder | 0.0603 | -0.372 | 0.00354 | 0.304 |
| U004 | Responder | 0.614 | -0.292 | -0.0834 | -0.225 |

Supplementary Table 10: **Average pre-DBS quadrant power across patients.** All pre-DBS power (a.u.) for each patient was averaged across 6-hour quadrants (0:00-06:00, 06:00-12:00, 12:00-18:00, 18:00-24:00).
